# Absence of nonfatal suicidal behavior preceding suicide death reveals differences in clinical risks

**DOI:** 10.1101/2024.06.05.24308493

**Authors:** Hilary Coon, Andrey Shabalin, Emily DiBlasi, Eric T. Monson, Seonggyun Han, Erin A. Kaufman, Danli Chen, Brent Kious, Nicolette Molina, Zhe Yu, Michael Staley, David K. Crockett, Sarah M. Colbert, Niamh Mullins, Amanda V. Bakian, Anna R. Docherty, Brooks Keeshin

## Abstract

Nonfatal suicidality is the most robust predictor of suicide death. However, only ∼10% of those who survive an attempt go on to die by suicide. Moreover, ∼50% of suicide deaths occur in the absence of prior known attempts, suggesting risks other than nonfatal suicide attempt need to be identified. We studied data from 4,000 population-ascertained suicide deaths and 26,191 population controls to improve understanding of risks leading to suicide death. This study included 2,253 suicide deaths and 3,375 controls with evidence of nonfatal suicidality (SUI_SI/SB and CTL_SI/SB) from diagnostic codes and natural language processing of electronic health records notes. Characteristics of these groups were compared to 1,669 suicides with no prior nonfatal SI/SB (SUI_None) and 22,816 controls with no lifetime suicidality (CTL_None). The SUI_None and CTL_None groups had fewer diagnoses and were older than SUI_SI/SB and CTL_SI/SB. Mental health diagnoses were far less common in both the SUI_None and CTL_None groups; mental health problems were less associated with suicide death than with presence of SI/SB. Physical health diagnoses were conversely more often associated with risk of suicide death than with presence of SI/SB. Pending replication, results indicate highly significant clinical differences among suicide deaths with versus without prior nonfatal SI/SB.

## 1. INTRODUCTION

The suicide death rate continues to rise precipitously in the U.S.^1^ Progress has been made in identifying risks leading to suicide attempts,^2-4^ but knowledge of risks specific to the outcome of suicide death remain largely elusive.^5-6^ At this time, nonfatal suicide attempt remains the strongest predictor of later suicide death,^7-9^ but few individuals who make suicide attempts go on to die by suicide. Estimates range from only 2.5% (three-year follow-up period)^10^ to 3.9% (five-year follow-up period)^11^ to 7% (nine-year follow-up period).^12^

The need to study risks specific to suicide mortality is therefore imperative. As an outcome, suicide death is distinct in many ways from suicide attempt. While suicide attempt is more than twice as likely in females than in males,^13^ suicide death is close to four times more frequent in men compared to women in the U.S. and many other countries.^14-15^ Recent results are also beginning to reveal genetic factors specific to risk of suicide death independent of genetic risks associated with psychopathology and/or suicide attempt.^16-18^ However, suicide deaths are comprised of two roughly equally sized subgroups: those who have evidence of nonfatal attempts, who may be more similar to living individuals with suicidality, and those with no prior nonfatal attempts.

Better understanding of clinical aspects of these subgroups will likely advance understanding of risks leading to suicide mortality. Co-occurring psychopathology has been broadly implicated in risk of nonfatal suicidal outcomes,^19^ and is often assumed to be a ubiquitous risk factor also for suicide death. However, evidence indicates that 80%-90% of individuals with psychiatric diagnoses will *not* go on to die by suicide,^20-21^ suggesting that psychiatric risk is unlikely to be a sufficient condition for risk of suicide death. While predictive models of suicide attempt and suicide death within populations of individuals with psychiatric diagnoses are reaching acceptable rates of risk prediction,^22-23^ studies within non-clinic samples reveal a more complex risk landscape that strongly implicates aspects of physical health and other risks in addition to psychiatric diagnoes.^24-26^ Recent work has further highlighted the multidimensional and transdiagnostic nature of risk of suicide death with modifiable risk factors found across disparate clinical and environmental domains including physical illness and sociodemographic and environmental factors.^27^ In a latent class analysis of suicide death in the National Violent Death Reporting System, the largest risk cluster exhibited physical health conditions with low prior nonfatal suicidality and little evidence of psychiatric risk.^28^ It is unknown the degree to which the difference in clinical risks represent underlying biological differences versus societal and care delivery issues (e.g., undiagnosed mental health conditions due to insufficient mental health screening, lack of access to mental health services, and/or lack of care-seeking behavior).

Our study compared the characteristics of suicide deaths with and without evidence of prior nonfatal suicidality through comprehensive characterization of the large, population-ascertained sample of individuals who died by suicide in the Utah Suicide Mortality Risk Study (USMRS), stratifying by presence versus absence of prior suicidal ideation and/or nonfatal suicidal behavior (SI/SB) documented in the healthcare system. Because SI/SB is not well captured with diagnoses in the electronic health records (EHR), we supplemented case and control ascertainment using data from Natural Language Processing (NLP) of free text health care notes.

## 2. METHODS

### 2.1. Sample

This study used data from a subset of 4,000 suicide deaths from the Utah Suicide Mortality Risk Study (USMRS) sample with available EHR notes in the University of Utah Health Sciences Center (UUHSC) system, one of two large health care providers in Utah. The USMRS resource has been described in detail elsewhere.^18^ Briefly, suicide determination was made by the centralized Utah State Office of the Medical Examiner (OME) following detailed investigation of the scene and circumstances of the death, and was given conservatively due to its impact on survivors. Deaths were securely linked to health data within the Utah Population Database (UPDB,)^29^ by UPDB staff. The UPDB is a state-wide database that contains over 27 million data records on over 12 million individuals, including demographic information and two decades of health records data. Data linking and NLP processing was done by UPDB staff and by honest data brokers. Identifiers were subsequently stripped, rare diagnoses and demographic groups were aggregated, and NLP results were collapsed into ratings of positive/negative suicidality before data were given to the research team to protect privacy and confidentiality. For each suicide death, records from ten living individuals in the Utah population with the same sex and birth year were matched using at-risk sampling. Only health records up to the time of matching to the index suicide death were studied, and 39 controls who died by suicide after matching were removed from the control group for this study. Because controls were matched using only sex and birth year, not all linked to health records resulting in 26,152 matched controls. Control data were similarly processed, aggregated, and stripped of identifiers prior to analysis by the research team. This study was approved by Institutional Review Boards from the University of Utah and the Utah Department of Health and Human Services.

### 2.2. Diagnostic data

Suicides and controls were defined as having nonfatal suicidality in part from diagnostic data through ICD-9 or ICD-10 diagnoses.^30,31^ The codes designating suicidal ideation or behavior (SI/SB) are listed in Supplemental Table S1. For suicide deaths, diagnoses that occurred within one week prior to death were excluded to eliminate diagnoses associated with the final fatal suicide event (cardiac arrest, respiratory failure, etc.) rather than those reflecting prior risk. For this analysis, ideation and behavior were combined into a single variable to be as conservative as possible in eliminating suicidality from the case and control groups assumed to have no SI/SB.

### 2.3. NLP from health care notes

The NLP was based on the criteria for suicidal behavior and ideation in the Columbia Suicide Severity Scale CSSRS,^32^ a questionnaire widely used in clinical practice and by agencies such as the FDA to assess the presence of SI/SB. The NLP is a rule-based algorithm detecting free text terms indicating current and lifetime suicidal behaviors and ideation. A version of this algorithm was validated in two large health care systems in the US and UK.^33^ The algorithm identifies both affirmative and negative mentions, and assigns a positive score if there is at least one affirmative mention. We manually validated the NLP in our Utah health records system on a pilot sample of notes from Utah suicide deaths and controls, finding a precision of 0.936 and an F1 score of 0.967 (see Supplemental tables S2-S7 for details). As with the diagnostic data, suicidal ideation and behavior from the NLP were combined.

### 2.4. Group definition and analyses using diagnostic codes and NLP

A suicide death or control was defined as positive for prior nonfatal suicidality (SI/SB) from relevant ICD-9/ICD-10 codes and from the NLP screened across all types of health care notes if at least one note was positive for current or historical ideation or behavior (SI/SB). This definition resulted in inclusive groups for SUI_SI/SB and CTL_SI/SB, with SUI_None and CTL_None being exclusive of documented prior suicidality. Importantly, as with the diagnostic ICD data, NLP from notes within one week prior to suicide death were excluded to eliminate mentions associated with the fatal attempt.

### 2.5. Other co-occurring conditions relating to mental health

Mental health diagnoses collapsed into interpretable groups using the hierarchical classification of diagnoses in the PheWAS catalog^34-35^ (PheCode Map 1.2 ICD10-CM), as implemented in the PheWAS R package.^36^ We began with the 1,866 detailed PheCodes, then retained 64 PheCodes associated with mental health, omitting those that substantially overlapped with diagnostic codes that defined the SI/SB groups in our study or that were overly general and difficult to interpret (Supplemental Table S8). Logistic regression was used wherein each PheCode was regressed on first on all suicides vs. all controls (suicide death effect, independent of presence of SI/SB) and then in separate models on all with SI/SB vs. all without SI/SB (independent of suicide or control status) to determine the relative independent impacts of suicide death versus presence of SI/SB. All tests adjusted for covariates detailed below. Results are reported as standardized marginal effects from the models.^37^ The adjusted significance threshold was 0.05/(2*64) = 3.91E-04. A mapping of each of the PheCodes to its specific ICD-9 and ICD-10 codes for all 36 PheCodes where either the effect of suicide death or the effect of presence of SI/SB met this significance threshold is given in Supplemental Table S9.

### 2.6. Additional analyses of PheCodes associated with physical health conditions

We studied 319 relatively common physical health PheCodes where each code had a prevalence of 5% or greater in any one of the four study groups (CTL_None, CTL_SI/SB, Sui_None, SUI_SI/SB). As with the mental health analysis, each PheCode was regressed on first on case/control status (suicide death effect) and then in separate models on presence of SI/SB, adjusting for covariates. The significance threshold was 0.05/(2*319) = 7.84E-05. Health conditions spanned multiple clinical areas.

### 2.7. Covariates for PheCode analyses

All analyses were adjusted for effects of age and sex. In addition, we observed significantly more diagnoses among SUI_SI/SB and CTL_SI/SB groups than the SUI_None and CTL_None groups. This effect introduces bias such that any co-occurring condition would systematically have more chance of being observed in the SUI_SI/SB and CTL_SI/SB groups (informative presence bias).^38-39^ For tests of specific conditions, we therefore additionally adjusted for overall number of ICD diagnoses, applying a square root transformation because of skewness in the distribution of observed number of diagnoses.

### 2.8. Sensitivity tests

Additional sensitivity tests assessed for: nonlinear effects of age to assure that results particularly for later onset diagnoses were not altered by more complex age effects; age x sex interaction effects to assure that results were not driven by more complex interactions; and year of death time-period cohort effect (defined in 5-year increments: <1998-2002, 2003-2007, 2008-2012, 2013-2017, 2018-2022) to assure that results were not altered by shifting trends in diagnostic practices. Within the suicide death groups, we tested for effects of violent method of death to assure that diagnostic effects were not substantively driven by associations with lethality. Violent method of death was defined as gun-related, hanging, cutting, or other violent trauma.

## 3. RESULTS

### 3.1. Defining presence/absence of nonfatal suicidality

Diagnoses within the UUHSC system linked to 4,000 Utah suicide deaths. Of these, 1669/4000=41.73% had evidence of prior nonfatal suicidality (SI/SB) from ICD-9 or ICD-10 codes. In addition, these deaths had 262,289 UUHSC health care notes across all types of encounters that were annotated by the NLP. Adding positive nonfatal suicidality from NLP to the determination from ICD codes resulted in 2253/4000=56.33% in the overall SUI_SI/SB group from all sources, an increase of 32% over and above the determination from ICD codes alone. Disagreement between determination of positive SI/SB from ICD codes vs. notes were primarily due to instances where there was a positive mention in notes but no corresponding positive ICD code (N=584). Of note, only 24/4000=0.6% of suicides had an ICD diagnostic code for suicidality, but no positive designation from the NLP, either due to missing note data (N=16) or false negative rating in the NLP (N=8).

Similarly, ICD data and NLP data were used to define control groups with SI/SB (CTL-SI/SB) and without SI/SB (CTL-None). There were 26,191 controls with diagnostic data in the UUHSC system. Of these, 1686/26191=6.44% were positive for SI/SB using ICD-9 or ICD-10 diagnostic codes. For these 26,191 controls, a total of 1,005,814 UUHSC notes across all types of encounters were annotated using the NLP. Adding positive controls for SI/SB using the NLP to the determination using ICD codes resulted in 3375/26191=12.89%, approximately doubling the positive rate over the determination from ICD codes alone. Disagreement in positive SI/SB determination was again predominantly driven by positive mentions in notes without corresponding ICD diagnoses positive for SI/SB (N=1,689). Instances where ICD was positive but the NLP was not positive occurred in 30 controls, with 22 of these due to false negative NLP results.

Table 1 shows basic descriptive characteristics of Utah suicide deaths and controls with SI/SB and without SI/SB. Significant differences of age and number of PheCodes were found between suicides and controls (independent of presence of SI/SB), and between all with SI/SB vs. all without SI/SB (independent of case status). All covariates were retained for subsequent analyses including effects of sex due to expected residual effects of sex on diagnoses associated with mental health.

**Table 1.** Tests of independent effects of age, sex, and number of diagnostic codes between suicide deaths and population controls with and without prior nonfatal suicide ideation or behavior (SI/SB), where SI/SB was determined from both diagnostic codes and NLP of EHR notes.

| Characteristic | CTL_None<br>(22,816) | CTL_SI/SB<br>(3,375) | All CTL<br>(26,191) | SUI_None<br>(1,747) | SUI_SI/SB<br>(2,253) | All SUI<br>(4,000) | Effect of suicide<br>death: OR (CI, p-value) | Effect of having<br>SI/SB: OR (CI, p-value) |
| --- | --- | --- | --- | --- | --- | --- | --- | --- |
| Mean age (std<br>dev) | 47.01<br>(17.71) | 40.31<br>(15.29) | 46.14<br>(17.56) | 47.73<br>(18.14) | 41.77<br>(15.07) | 44.35<br>(16.73) | 0.988 (0.986-0.990,<br><0.0001) | 0.961 (0.959-0.963,<br><0.0001) |
| Percent male<sup>1</sup> | 73.30% | 69.96% | 72.86% | 80.53% | 63.31% | 70.76% | 1.05 (0.97-1.13, NS) | 1.02 (0.95-1.10, NS) |
| Mean overall N<br>PheCodes (std<br>dev)<sup>2</sup> | 47.27<br>(48.94) | 96.59<br>(89.28) | 53.65<br>(58.23) | 51.81<br>(49.01) | 95.74<br>(90.48) | 76.75<br>(78.47) | 1.123 (1.113-1.134,<br><0.0001) | 1.324 (1.311-1.337,<br><0.0001) |

<sup>1</sup> Coded male=1, female=0 for the logistic regression test.
<sup>2</sup> Raw N of codes is presented; for statistical testing, this variable was square root transformed because of skewness.

### 3.2. Diagnostic comparisons of mental health conditions

Of the 64 PheCodes included, 32 were significant using the adjusted threshold of 3.915E-04 for either the effect of suicide death or the presence vs. absence of SI/SB effects (Table 2, Figure 1). Of the 32 other mental health PheCodes without significant effects, 27 were relatively rare in our study sample, with <5% prevalence across all four of the comparison groups (SUI_SI/SB, SUI_None, CTL_SI/SB, and CTL_None).

**Table 2.** Standardized marginal effects of logistic regressions of presence/absence of mental health PheCodes PheCodes on case-control status (suicide death effect) and also on presence/absence of SI/SB (presence of SI/SB effect).

| PheCode | Grouping | Effect: suicide death (p-value) | Effect: presence SI/SB (p-value) |
| --- | --- | --- | --- |
| Depression | Affective | **0.938 (8.32E-85)** | **2.134 (<E-350)** |
| Bipolar | Affective | **0.793 (3.31E-42)** | **2.119 (1.47E-283)** |
| Major Depressive Disorder | Affective | **0.621 (3.34E-35)** | **2.404 (<E-350)** |
| Dysthymic disorders | Affective | **0.446 (3.50E-11)** | **1.252 (6.96E-81)** |
| Other mood disorders | Affective | **0.367 (4.76E-08)** | **2.007 (2.26E-193)** |
| Somatoform disorder | Anxiety/stress | **0.867 (9.54E-14)** | **0.888 (1.96E-12)** |
| Anxiety-related states & conditions | Anxiety/stress | **0.787 (7.03E-35)** | **2.700 (8.18E-304)** |
| Posttraumatic stress disorder | Anxiety/stress | **0.736 (5.29E-24)** | **1.914 (9.40E-132)** |
| Anxiety disorder | Anxiety/stress | **0.709 (1.78E-52)** | **1.388 (1.13E-265)** |
| Agoraphobia, social phobia, panic disorder | Anxiety/stress | **0.531 (3.76E-16)** | **1.520 (1.50E-122)** |
| Obsessive-compulsive disorders | Anxiety/stress | **0.404 (8.77E-05)** | **1.701 (2.11E-53)** |
| Generalized anxiety disorder | Anxiety/stress | **0.356 (4.08E-10)** | **1.611 (2.34E-203)** |
| Eating disorders | Anxiety/stress | 0.481 (0.015) | **1.517 (3.27E-11)** |
| Acute reaction to stress | Anxiety/stress | 0.268 (0.0069) | **0.757 (4.20E-15)** |
| Adjustment reaction | Anxiety/stress | 0.243 (0.0027) | **1.232 (1.77E-54)** |
| Personality disorders (excluding antisocial PD and borderline PD) | PD | **0.961 (2.72E-28)** | **2.362 (5.20E-109)** |
| Antisocial PD & borderline PD | PD | **0.884 (8.66E-23)** | **2.407 (2.29E-101)** |
| Conduct disorders | Impulsive | 0.181 (0.072) | **1.716 (6.02E-65)** |
| Attention deficit hyperactivity disorder | Impulsive | 0.180 (0.0067) | **1.158 (2.59E-81)** |
| Developmental delays & disorders | Developmental | 0.072 (0.301) | **0.990 (7.70E-56)** |
| Delirium | Psychotic | **0.863 (1.10E-18)** | **1.013 (8.77E-23)** |
| Alteration of consciousness | Psychotic | **0.781 (2.55E-30)** | **0.883 (9.57E-39)** |
| Paranoid disorders | Psychotic | **0.733 (5.77E-13)** | **2.186 (4.87E-73)** |
| Psychosis | Psychotic | **0.685 (1.77E-23)** | **1.822 (5.65E-142)** |
| Schizophrenia | Psychotic | **0.643 (9.66E-15)** | **2.339 (1.15E-134)** |
| Altered mental status | Psychotic | **0.621 (2.88E-17)** | **0.888 (2.19E-35)** |
| Hallucinations | Psychotic | 0.381 (0.0014) | **1.980 (3.89E-48)** |
| Substance addiction & disorders | SUD | **1.100 (1.95E-113)** | **1.644 (2.51E-308)** |
| Alcoholism | SUD | **1.018 (6.01E-94)** | **1.425 (1.96E-197)** |
| Alcohol-related disorders | SUD | **1.013 (4.06E-68)** | **1.652 (2.13E-170)** |
| Adverse events of opiates & narcotics in therapeutic use | SUD | **0.744 (2.54E-24)** | **0.705 (7.75E-23)** |
| Tobacco use disorder | SUD | **0.718 (6.96E-67)** | **0.742 (6.97E-85)** |

Notes. Standardized marginal effects were derived from logistic regressions.<sup>37</sup> Each binary presence/absence PheCode was regressed on case-control status (suicide death effect) and then on presence/absence of SI/SB (presence SI/SB effect), adjusting for age, sex, and transformed overall number of PheCodes. Tests that were significant after adjustment for multiple testing are shown in bold type.

**Figure 1.**
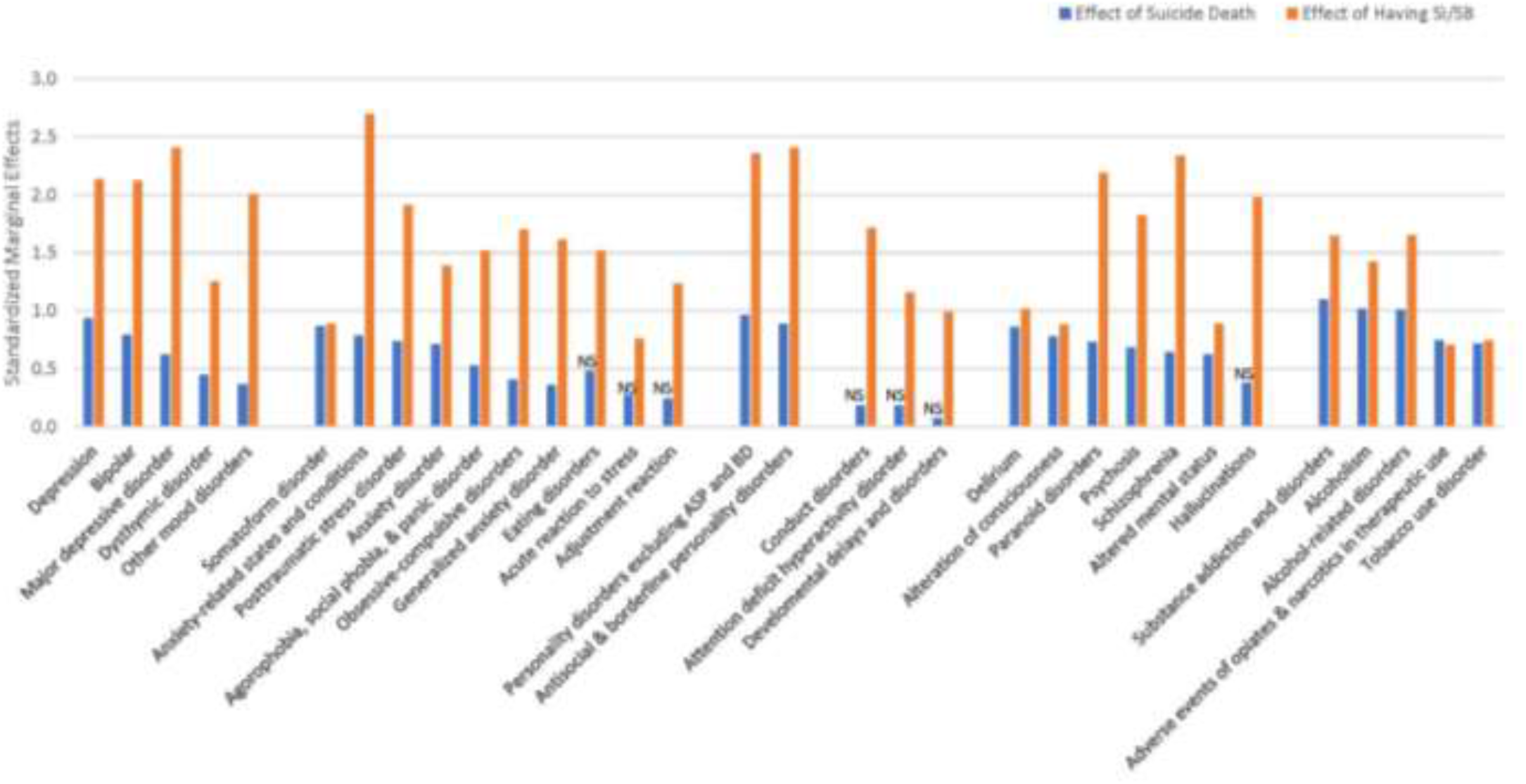
Standardized margnal effects of logistic regressions of presence/absence of mental health PheCodes on case-control status (blue: suicide death effect) and also on presence/absence of Sl/58 (fed: have SI/SB effect). Note. Standardized marginal effects were derived from logistic regressions ^37^ Each binary presence/absence PheCode was regressed on case-control status (blue: suicide death effect) and on presence/absence of Sl/SB (red: have Sl/SB effect), adjusting for age, sex, and transformed overall number of PheCodes.

Across the 32 significant mental health PheCodes, presence of mental health diagnoses was much more strongly associated with presence/absence of SI/SB than with suicide death (Table 2). Indeed, on average, presence of SI/SB on psychiatric diagnoses had statistical marginal effect sizes over three times greater than the effects of suicide death (Table 2 and Figure 1). The difference in the associations of SI/SB vs. suicide death was smallest for substance use disorders, and greatest for developmental disorders, conduct disorder, and ADHD.

Supplemental Figure 1 shows the prevalence of PheCodes within each subgroup (CTL_None, CTL_SI/SB, SUI_None, SUI_SI/SB) adjusted for effects of age, sex, and transformed overall number of diagnoses, further illustrating the differences in associations of suicide death vs. associations of presence of SI/SB. The figure shows SUI_SI/SB is generally more similar to CTL_SI/SB than to SUI_None, with exceptions for tobacco use disorder and adverse effects of opioids.

### 3.3. Characteristics involving other physical health conditions

Of the 319 physical health PheCodes, 133 were significantly associated with either suicide death or presence of SI/SB. Among these, 82 showed significant effects only in the negative (protective) direction for either suicide death or presence of SI/SB (Supplemental Table S10). While these PheCodes are of interest, they will be the focus of future studies, as this analysis is focused on aspects leading to suicide risk. The results presented in Table 3 and Figure 2 therefore reflect 51 PheCodes with significant positive (risk) effects associated with either suicide death or presence of SI/SB.

**Table 3.** Standardized marginal effects of logistic regressions of presence/absence of physical health PheCodes PheCodes on case-control status (suicide death effect) and also on presence/absence of SI/SB (presence of SI/SB effect).

| PheCode | Grouping | Effect: suicide death (p-value) | Effect: presence SI/SB (p-value) |
| --- | --- | --- | --- |
| Tachycardia NOS | Circulatory | **0.274 (1.11E-05)** | 0.209 (2.20E-04) |
| Acute pancreatitis | Digestive | **0.562 (2.35E-08)** | 0.140 (0.161) |
| Peptic ulcer | Digestive | **0.517 (6.72E-10)** | 0.253 (0.0022) |
| Hematemesis | Digestive | **0.486 (4.29E-05)** | 0.352 (0.0027) |
| Esophagitis, GERD, and related diseases | Digestive | **0.298 (7.36E-05)** | -0.187 (0.0087) |
| Diseases of teeth and supporting structures | Digestive | 0.069 (0.437) | **0.453 (2.15E-08)** |
| Acidosis | Endocrine | **0.605 (8.59E-16)** | **0.366 (4.69E-07)** |
| Hypopotassemia | Endocrine | **0.570 (2.48E-20)** | **0.348 (2.41E-09)** |
| Protein-calorie malnutrition | Endocrine | **0.428 (7.01E-07)** | 0.224 (0.0070) |
| Hypovolemia | Endocrine | **0.390 (3.23E-14)** | 0.052 (0.257) |
| Acute renal failure | Genitourinary | **0.425 (1.25E-08)** | 0.082 (0.257) |
| Renal dialysis | Genitourinary | 0.329 (0.0033) | **0.594 (2.52E-09)** |
| Urinary tract infection | Genitourinary | **0.239 (2.39E-05)** | -0.164 (0.0015) |
| Viral hepatitis C | Infectious | 0.305 (0.0047) | **0.955 (2.33E-19)** |
| Crushing or internal injury to organs | Injuries | **0.607 (4.19E-11)** | -0.106 (0.238) |
| Adverse drug events or drug allergies | Injuries | **0.575 (2.66E-20)** | **0.681 (1.37E-31)** |
| Fracture of ribs | Injuries | **0.481 (3.01E-08)** | 0.090 (0.291) |
| Fracture of vertebral column w/o spinal cord injury | Injuries | **0.463 (5.14E-06)** | -0.091 (0.277) |
| Open wounds of extremities | Injuries | **0.456 (1.39E-17)** | **0.379 (3.19E-14)** |
| Concussion | Injuries | **0.439 (1.99E-10)** | -0.016 (0.807) |
| Complication of internal orthopedic device | Injuries | **0.364 (2.62E-05)** | **-0.391 (2.99E-06)** |
| Sprains and strains of back and neck | Injuries | **0.359 (1.72E-14)** | 0.140 (0.00104) |
| Open wound of hand | Injuries | **0.335 (2.32E-06)** | 0.045 (0.498) |
| Burns | Injuries | **0.330 (4.76E-05)** | -0.040 (0.596) |
| Superficial injury w/o infection | Injuries | **0.328 (1.36E-11)** | **0.183 (3.86E-05)** |
| Contusion | Injuries | **0.313 (1.88E-13)** | 0.077 (0.0456) |
| Open wound of head and face | Injuries | **0.281 (1.27E-07)** | 0.061 (0.211) |
| Skull and face fracture and other intercranial injury | Injuries | **0.270 (2.22E-05)** | **0.288 (9.39E-05)** |
| Personal history of medication allergy | Injuries | 0.223 (0.0184) | **0.665 (1.18E-13)** |
| Injury, NOS | Injuries | **0.205 (2.36E-06)** | 0.035 (0.366) |
| Other and unspecified disc disorder | Musculoskeletal | **0.469 (3.88E-08)** | -0.266 (0.0015) |
| Degeneration of intervertebral disc | Musculoskeletal | **0.277 (6.00E-07)** | **-0.231 (1.15E-05)** |
| Spondylosis w/o myelopathy | Musculoskeletal | **0.252 (2.70E-05)** | **-0.283 (7.89E-07)** |
| Displacement of intervertebral disc | Musculoskeletal | **0.248 (4.76E-05)** | -0.212 (2.63E-04) |
| Chronic pain syndrome | Neurologic | **0.755 (5.09E-12)** | **0.504 (6.23E-06)** |
| Convulsions | Neurologic | **0.539 (1.86E-16)** | **0.308 (6.82E-07)** |
| Encephalopathy | Neurologic | **0.462 (1.16E-06)** | **0.515 (2.59E-08)** |
| Unspecified disorders of nervous system | Neurologic | **0.460 (9.20E-06)** | **0.425 (2.71E-05)** |
| Coma | Neurologic | **0.442 (8.14E-05)** | **0.777 (5.78E-12)** |
| Insomnia | Neurologic | **0.366 (1.64E-12)** | **0.928 (1.92E-88)** |
| Chronic pain conditions | Neurologic | **0.354 (3.27E-10)** | **0.404 (2.26E-15)** |
| Abnormal involuntary movements | Neurologic | **0.336 (1.44E-05)** | **0.361 (8.44E-07)** |
| Migraine | Neurologic | **0.297 (1.49E-06)** | 0.188 (8.22E-04) |
| Other sleep disorders | Neurologic | -0.173 (0.0363) | **0.351 (1.52E-06)** |
| Pneumonitis due to inhalation of food or vomit | Respiratory | **0.968 (5.14E-25)** | **0.780 (2.10E-15)** |
| Respiratory failure | Respiratory | **0.737 (3.50E-23)** | 0.250 (6.11E-04) |
| Chronic airway obstruction | Respiratory | **0.481 (9.79E-08)** | 0.103 (0.259) |
| Asphyxia and hypoxemia | Respiratory | **0.277 (4.77E-05)** | -0.019 (0.767) |
| Rhabdomyolysis | Symptoms<sup>1</sup> | **0.916 (3.10E-13)** | **1.036 (3.51E-14)** |
| Syncope and collapse | Symptoms | **0.250 (2.21E-05)** | 0.099 (0.0699) |
| Cervicalgia | Symptoms | **0.246 (3.15E-06)** | -0.048 (0.322) |

1 Symptoms is a PheWas category including symptoms resulting from conditions across multiple clinical domains
Notes. Standardized marginal effects were derived from logistic regressions.<sup>37</sup> Each binary presence/absence PheCode was regressed on case-control status (suicide death effect) and then on presence/absence of SI/SB (presence SI/SB effect), adjusting for age, sex, and transformed overall number of PheCodes. Tests that were significant after adjustment for multiple testing are shown in bold type.

**Figure 2.**
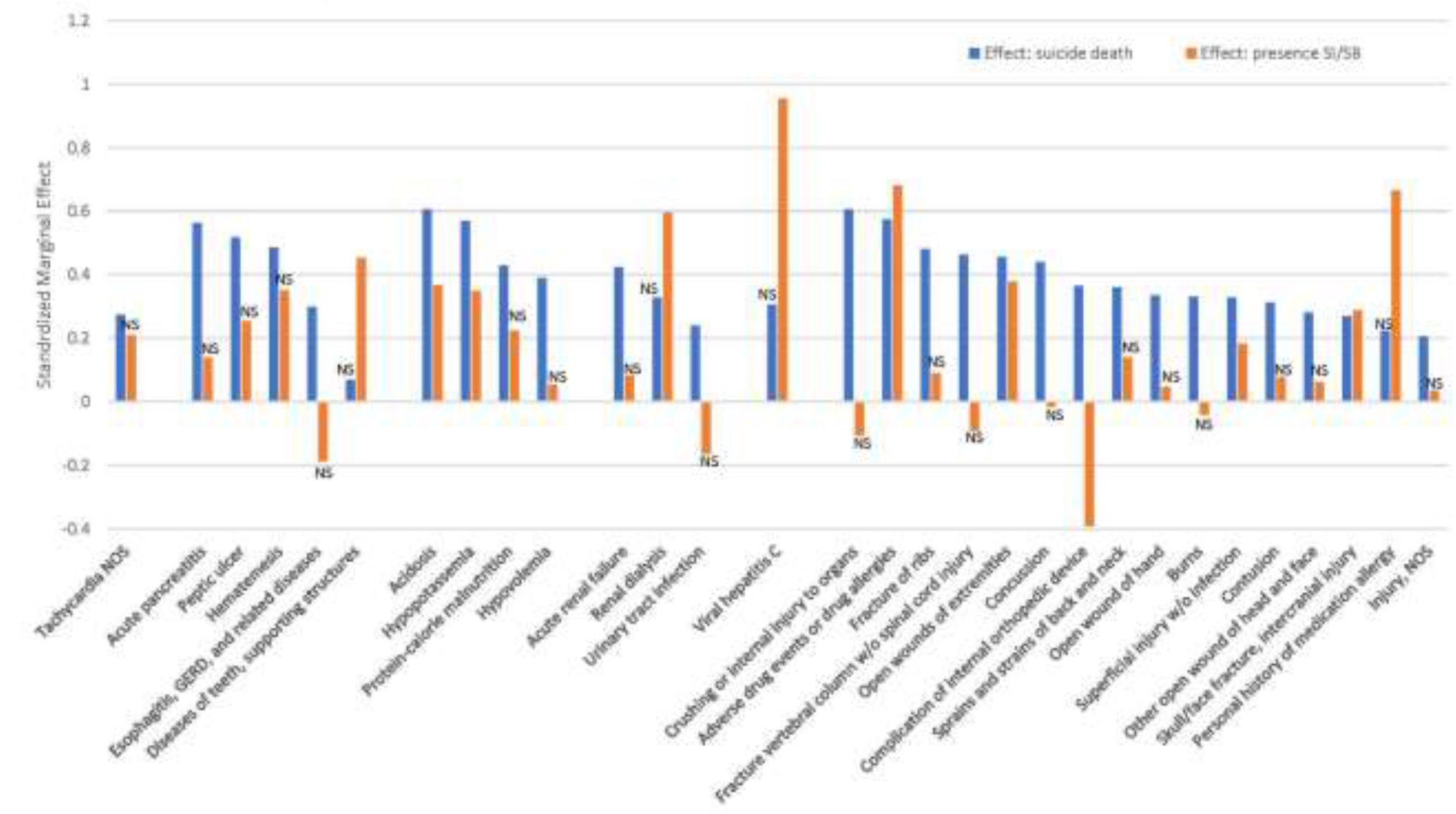
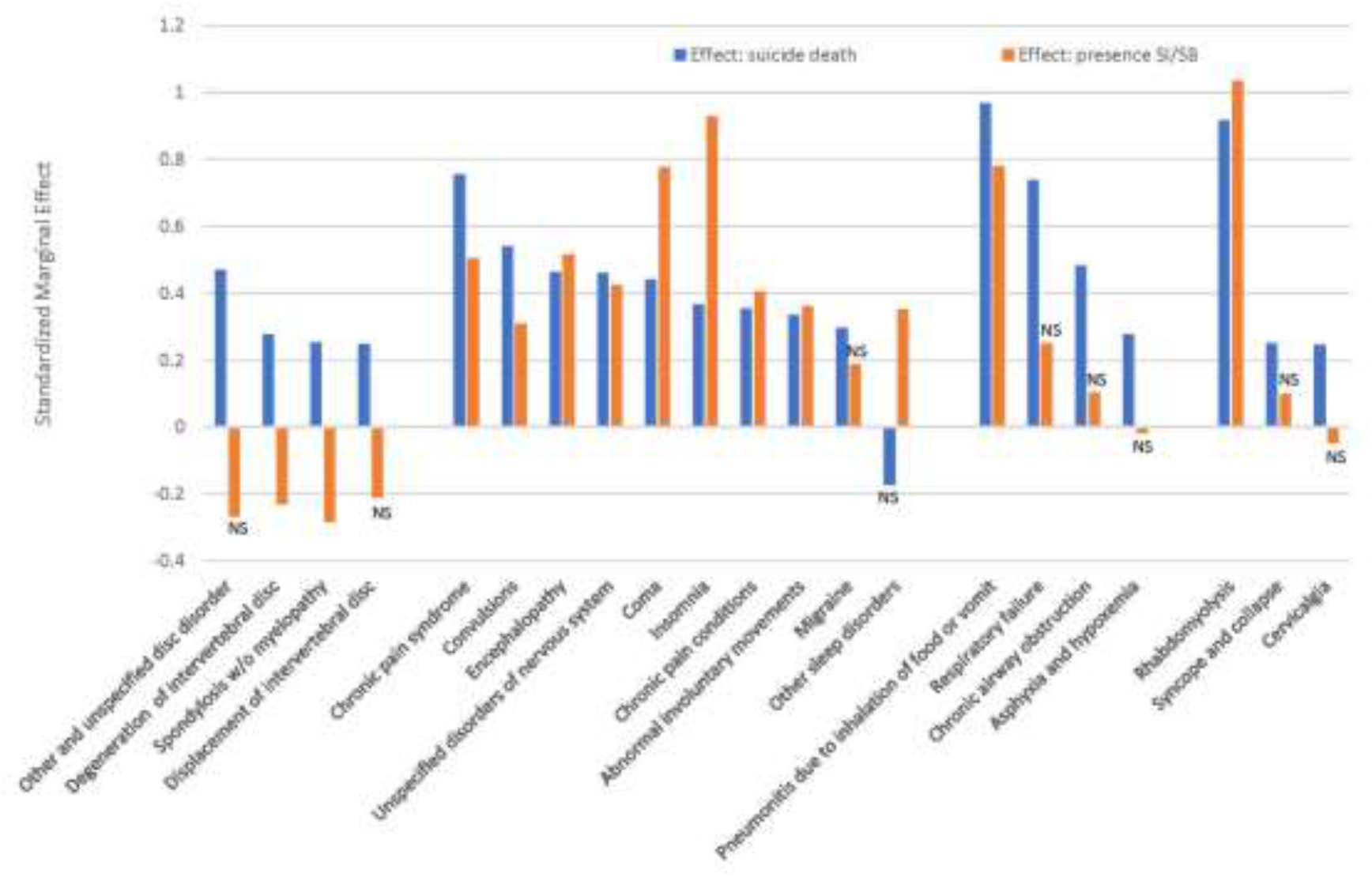
Standardised marginal effects of logistic regressions of presence/absence of physical health PheCodes on case-control status (blue, suicide death effect) and also on presence/absence of SI/S9 (red: have SI/VSB effect) Note. Standardized marginal effects were derived from logistic regressions ^37^ Each binary presence/absence PheCode was regressed on case-control status (blue: suicide death effect) and on presence/absence of Sl/SB (red: have Sl/SB effect), adjusting for age, sex, and transformed overall number of PheCodes.

In contrast to the mental health conditions, these PheCode results showed on average larger marginal standardized effects associated with suicide death than with presence of SI/SB (Figure 2). For 27 of the 51 PheCodes, there was a significant association with suicide death but no significant association with presence of SI/SB. An additional 10 of the PheCodes showed significant but smaller associations with SI/SB than with suicide death, and five PheCodes had approximately equivalent effects. Results also included three conditions with opposing significant results: positive (risk) associations with suicide death and negative (protective) associations with presence of SI/SB. Only six of the physical health PheCodes showed stronger associations with SI/SB than suicide death.

### 3.4. Sensitivity tests of non-linear age effects and age by sex interactions

Main analyses used a parsimonious adjustment for age, sex, and transformed overall N of diagnoses. For the results in Tables 2 and 3, we additionally applied sensitivity tests for nonlinear effects of age, for age by sex interactions, and for time period cohort effects. Inclusion of these additional covariates did not alter significance status of main effects, and the prevalence (adjusted for covariates) of mental health-associated PheCodes changed only modestly, resulting in average changes of 0.005, 0.004, 0.003, and 0.001 for CTL_None, CTL_SI/SB, SUI_None, and SUI_SI/SB, respectively (Supplemental Table S11). Results were similar for PheCodes associated with physical conditions, again not altering overall main effects, and resulting in only modest changes in prevalences (0.008, 0.007, 0.005, and 0.003, for CTL_None, CTL_SI/SB, SUI_None, and SUI_SI/SB, respectively; Supplemental Table S12).

### 3.5. Violent vs. non-violent method of suicide death

Sensitivity tests of violent suicide death (adjusting for age, sex, and transformed N of diagnoses) were limited to the 2 suicide death groups. The effect of violent death on prevalence of mental health PheCodes across the four groups was not significant adjusting for multiple testing, with a few exceptions, including significantly decreased prevalence of Depression, Anxiety-related conditions, Substance addiction and disorders, and Opioid adverse events (Supplemental Table S13). These observed changes were independent of occurrence of SI/SB and did not affect overall substantive results. No physical health PheCodes had significant effects of violent death after adjusting for multiple testing.

## 4. DISCUSSION

This study provides characterization of a large sample (N=4000) of suicide deaths with evidence of prior nonfatal suicidal ideation or behavior (SI/SB) versus those with no prior evidence of SI/SB, comparing to data from 26,152 matched controls with and without SI/SB. Evidence of nonfatal suicidal ideation or behaviors (SUI_SI/SB and CTL_SI/SB) came from sources including diagnostic codes supplemented by information from NLP applied to notes in the EHR. Defining groups for presence/absence of SI/SB using both diagnoses and the NLP resulted in more accurate determination; the NLP increased the SUI_SI/SB group size by about 35%, and the CTL_SI/SB group size by about 50%. The performance of this NLP suggests the importance of not relying solely on the ICD diagnostic system to identify those with positive suicidality. Importantly, the defined groups without SI/SB (SUI_None and CTL_None) were less likely to have false negative instances of undetected SI/SB than if SI/SB determination had only been based on ICD diagnostic codes.

Demographic comparisons of the four resulting groups showed that the SUI_SI/SB and CTL_SI/SB groups both were significantly younger at death (or matching age) than SUI_None and CTL_None (p<0.0001), and both had significantly more diagnoses across all PheCodes (p<0.0001), replicating previous results.^40^ We adjusted subsequent tests for this significant systematic difference in diagnoses following recommendations as described elsewhere.^38-39^

Tests of clinical diagnoses used PheCodes from the PheWAS hierarchical classification system,^34-35^ allowing us to collapse similar codes into interpretable groups. Thirty-two mental health related PheCodes were highly significantly increased in both the suicide and control groups with SI/SB. The association between presence of SI/SB and psychiatric diagnoses was on average over three times greater than the association with suicide death. Prevalences of PheCodes were strikingly similar across SUI_SI/SB and CTL_SI/SB for most psychiatric diagnoses; presence of SI/SB appears to be much more substantially associated with psychopathology than with suicide death. Sensitivity tests of non-linear effects of age, interactions, a time-period cohort effect, and violent method of death did not substantively alter these results.

Tests of physical health PheCodes resulted in a markedly different pattern, with significant effects associated with suicide death that were, on average, larger than effects associated with presence of SI/SB. These results replicate a large recent latent class study of suicide death.^28^ PheCodes with markedly higher suicide death associations than presence of SI/SB included those with challenging chronic pain and/or disability (spondylosis, degeneration or displacement of intervertebral discs, complications from an internal orthopedic device). In general, the conditions significant for suicide death were enriched for diagnoses associated with injuries and neurologic conditions associated with pain. While some of these may represent unreported suicide attempts (e.g., Open wounds of extremities, Adverse drug events or drug allergies), those with strong suicide death associations may have clinical relevance in the implication of pain as a risk factor leading to suicide mortality, even in the absence of documented SI/SB.

## 4.1. Limitations

The differences in clinical data specifically associated with mental health could result from less care-seeking behavior in the SUI_None group, which could stem from lack of access to mental health care services or social stigma, issues we are unable to measure in this study. We did attempt to minimize this possible effect by including data only from individuals with evidence of encounters with the health care system, and by adjusting all tests for the number of diagnostic codes in the records. Additionally, our data resource requires that SI/SB is defined by the administrative data available in the EHR. Though we attempted to minimize this effect through our addition of SI/SB from NLP in EHR notes, some of those in the SUI_None and CTL _None groups were likely experiencing undocumented nonfatal SI/SB. Finally, we note that while our data represent state-wide population ascertainment, results may not be generalizable to other populations.

### 4.2. Conclusions

This study uses a population-ascertained resource of suicide deaths and age-/birth year-matched control data to begin to address suicide mortality knowledge gaps. We augmented the definition of presence of SI/SB using both diagnoses and NLP of health notes to provide more inclusive groups with SI/SB and groups without SI/SB that are less likely to include false negatives. Results suggested powerful associations of presence of nonfatal SI/SB with psychopathology, with effects far overshadowing associations between psychopathology and suicide death. Results additionally implicated other physical conditions, particularly those associated with injury and pain, as potentially having specific relevance for risk of suicide death in the absence of documented prior nonfatal SI/SB. In addition to replication of these results, more definitive tests for additional underlying biological vulnerabilities that could partially drive these observed clinical results will require further study.

## Data Availability

All data produced in the present work are contained in the manuscript

## DECLARATION OF GENERATIVE AI AND AI-ASSISTED TECHNOLOGIES IN THE WRITING PROCESS

No AI tools were used in the writing of this manuscript.

## ACKNOWLEDGMENTS

This work was supported by the National Institute of Health (HC, grant number R01MH122412, R01MH123489; AD, grant number R01MH123619; AVB, grant number R01ES032028); the Brain & Behavior Research Foundation--NARSAD (EM, grant number 31248). Partial support for all datasets housed within the Utah Population Data Base is provided by the Huntsman Cancer Institute (HCI), http://www.huntsmancancer.org/, and the HCI Cancer Center Support grant, P30CA42014 from the National Cancer Institute. Research was supported by NCRR grant “Sharing statewide health data for genetic research” R01RR021746 with additional support from the Utah Department of Health and Human Services and the University of Utah. We thank University of Utah Health Data Science Services for data and analytics support, and the University of Utah Pedigree and Population Resource and the University of Utah Health Enterprise Data Warehouse for establishing the Master Subject Index between the Utah Population Database and the University of Utah Health Sciences Center. We thank Dr. Hunter Strohmeyer and Dr. Michael Pope for their assistance in manual validation of the NLP pipeline. We thank Drs. Marika Cusick, John Mann, and Jyotishman Pathak for their collaborative help in deploying the NLP pipeline and additional input on this study.

