## supplemental tables and figure for "Absence of nonfatal suicidal behavior preceding suicide death reveals differences in clinical risks"

**Supplemental Table S1. ICD-9 and ICD-10 diagnostic codes defining suicidal ideation and suicidal behavior**.

| Version | Code | Definition |
|---|---|---|
| ICD-9 | E950.0 | Suicide and self-inflicted poisoning by analgesics, antipyretics, and antirheumatics |
| ICD-9 | E950.1 | Suicide and self-inflicted poisoning by barbiturates |
| ICD-9 | E950.2 | Suicide and self-inflicted poisoning by other sedatives and hypnotics |
| ICD-9 | E950.3 | Suicide and self-inflicted poisoning by tranquilizers and other psychotropic agents |
| ICD-9 | E950.4 | Suicide and self-inflicted poisoning by other specified drugs and medicinal substances |
| ICD-9 | E950.5 | Suicide and self-inflicted poisoning by unspecified drug or medicinal substance |
| ICD-9 | E950.6 | Suicide and self-inflicted poisoning by agricultural and horticultural chemical and pharmaceutical preparations other than plant foods and fertilizers |
| ICD-9 | E950.7 | Suicide and self-inflicted poisoning by corrosive and caustic substances |
| ICD-9 | E950.8 | Suicide and self-inflicted poisoning by arsenic and its compounds |
| ICD-9 | E950.9 | Suicide and self-inflicted poisoning by other and unspecified solid and liquid substances |
| ICD-9 | E951.0 | Suicide and self-inflicted poisoning by gas distributed by pipeline |
| ICD-9 | E951.1 | Suicide and self-inflicted poisoning by liquefied petroleum gas distributed in mobile containers |
| ICD-9 | E951.8 | Suicide and self-inflicted poisoning by other utility gas |
| ICD-9 | E952.0 | Suicide and self-inflicted poisoning by motor vehicle exhaust gas |
| ICD-9 | E952.1 | Suicide and self-inflicted poisoning by other carbon monoxide |
| ICD-9 | E952.8 | Suicide and self-inflicted poisoning by other specified gases and vapors |
| ICD-9 | E952.9 | Suicide and self-inflicted poisoning by unspecified gases and vapors |
| ICD-9 | E953.0 | Suicide and self-inflicted poisoning by hanging |
| ICD-9 | E953.1 | Suicide and self-inflicted injury by suffocation by plastic bag |
| ICD-9 | E953.8 | Suicide and self-inflicted injury by other specified hanging, strangulation, and suffocation |
| ICD-9 | E953.9 | Suicide and self-inflicted injury by other unspecified hanging, strangulation, and suffocation |
| ICD-9 | E954 | Suicide and self-inflicted injury by submersion [drowning] |
| ICD-9 | E955.0 | Suicide and self-inflicted injury by handgun |
| ICD-9 | E955.1 | Suicide and self-inflicted injury by shotgun |
| ICD-9 | E955.2 | Suicide and self-inflicted injury by hunting rifle |
| ICD-9 | E955.3 | Suicide and self-inflicted injury by military firearms |
| ICD-9 | E955.4 | Suicide and self-inflicted injury by other and unspecified firearm |
| ICD-9 | E955.5 | Suicide and self-inflicted injury by explosives |
| ICD-9 | E955.6 | Suicide and self-inflicted injury by airgun |
| ICD-9 | E955.7 | Suicide and self-inflicted injury by paintball gun |
| ICD-9 | E955.9 | Suicide and self-inflicted injury by unspecified firearms, airguns, and explosives |
| ICD-9 | E956 | Suicide and self-inflicted injury by cutting and piercing instrument |
| ICD-9 | E957.0 | Suicide and self-inflicted injury by jumping from high place, residential premises |
| ICD-9 | E957.1 | Suicide and self-inflicted injury by jumping from high place, other man-made structures |
| ICD-9 | E957.2 | Suicide and self-inflicted injury by jumping from high place, natural sites |
| ICD-9 | E959.2 | Suicide and self-inflicted injury by jumping from high place, unspecified |
| ICD-9 | E958.0 | Suicide and self-inflicted injury by jumping or lying before moving object |
| ICD-9 | E958.1 | Suicide and self-inflicted injury by burns, fire |
| ICD-9 | E958.2 | Suicide and self-inflicted injury by scald |
| ICD-9 | E958.3 | Suicide and self-inflicted injury by extremes of cold |
| ICD-9 | E958.4 | Suicide and self-inflicted injury by electrocution |
| ICD-9 | E958.5 | Suicide and self-inflicted injury by crashing of motor vehicle |

| | | |
|---|---|---|
| ICD-9 | E958.6 | Suicide and self-inflicted injury by crashing of aircraft |
| ICD-9 | E958.7 | Suicide and self-inflicted injury by caustic substances, except poisoning |
| ICD-9 | E958.8 | Suicide and self-inflicted injury by other specified means |
| ICD-9 | E958.9 | Suicide and self-inflicted injury by unspecified means |
| ICD-9 | E959 | Late effects of self-inflicted injury |
| ICD-9 | V62.84 | Suicidal ideation |
| ICD10 | X71.0xx | Intentional self-harm by drowning and submersion while in bathtub |
| ICD10 | X71.1xx | Intentional self-harm by drowning and submersion while in swimming pool |
| ICD10 | X71.2xx | Intentional self-harm by drowning and submersion after jumping into swimming pool |
| ICD10 | X71.3xx | Intentional self-harm by drowning and submersion in natural water |
| ICD10 | X71.8xx | Other intentional self-harm by drowning and submersion |
| ICD10 | X71.9xx | Intentional self-harm by drowning and submersion, unspecified |
| ICD10 | X72.xxx | Intentional self-harm by handgun discharge |
| ICD10 | X73.0xx | Intentional self-harm by shotgun discharge |
| ICD10 | X73.1xx | Intentional self-harm by hunting rifle discharge |
| ICD10 | X73.2xx | Intentional self-harm by machine gun discharge |
| ICD10 | X73.8xx | Intentional self-harm by other larger firearm discharge |
| ICD10 | X73.9xx | Intentional self-harm by unspecified larger firearm discharge |
| ICD10 | X74.01x | Intentional self-harm by airgun |
| ICD10 | X74.02x | Intentional self-harm by paintball gun |
| ICD10 | X74.09x | Intentional self-harm by other gas, air, or spring-operated gun |
| ICD10 | X74.8xx | Intentional self-harm by other firearm discharge |
| ICD10 | X74.9xx | Intentional self-harm by unspecified firearm discharge |
| ICD10 | X75.xxx | Intentional self-harm by explosive material |
| ICD10 | X76.xxx | Intentional self-harm by smoke, fire, and flames |
| ICD10 | X77.0xx | Intentional self-harm by steam or hot vapors |
| ICD10 | X77.1xx | Intentional self-harm by hot tap water |
| ICD10 | X77.2xx | Intentional self-harm by other hot fluids |
| ICD10 | X77.3xx | Intentional self-harm by hot household appliances |
| ICD10 | X77.8xx | Intentional self-harm by other hot objects |
| ICD10 | X77.9xx | Intentional self-harm by unspecified hot objects |
| ICD10 | X78.0xx | Intentional self-harm by sharp glass |
| ICD10 | X78.1xx | Intentional self-harm by knife |
| ICD10 | X78.2xx | Intentional self-harm by sword or dagger |
| ICD10 | X78.8xx | Intentional self-harm by other sharp object |
| ICD10 | X78.9xx | Intentional self-harm by unspecified sharp object |
| ICD10 | X79.xxx | Intentional self-harm by blunt object |
| ICD10 | X80.xxx | Intentional self-harm by jumping from a high place |
| ICD10 | X81.0xx | Intentional self-harm by jumping or lying in front of motor vehicle |
| ICD10 | X81.1xx | Intentional self-harm by jumping or lying in front of (subway) train |
| ICD10 | X81.8xx | Intentional self-harm by jumping or lying in front of other moving object |
| ICD10 | X82.0xx | Intentional collision of motor vehicle with other motor vehicle |
| ICD10 | X82.1xx | Intentional collision of motor vehicle with train |
| ICD10 | X82.2xx | Intentional collision of motor vehicle with tree |
| ICD10 | X82.8xx | Other intentional self-harm by crashing of motor vehicle |
| ICD10 | X83.0xx | Intentional self-harm by crashing of aircraft |
| ICD10 | X83.1xx | Intentional self-harm by electrocution |
| ICD10 | X83.2xx | Intentional self-harm by exposure to extremes of cold |

| | | |
|---|---|---|
| ICD10 | X83.8xx | Intentional self-harm by other specified means |
| ICD10 | T36.0x2 | Poisoning by penicillins, intentional self-harm |
| ICD10 | T36.1x2 | Poisoning by cephalosporins and other beta-lactam antibiotics, intentional self-harm |
| ICD10 | T36.2x2 | Poisoning by chloramphenicol group, intentional self-harm |
| ICD10 | T36.3x2 | Poisoning by macrolides, intentional self-harm |
| ICD10 | T36.4x2 | Poisoning by tetracyclines, intentional self-harm |
| ICD10 | T36.5x2 | Poisoning by aminoglycosides, intentional self-harm |
| ICD10 | T36.6x2 | Poisoning by rifampicins, intentional self-harm |
| ICD10 | T36.7x2 | Poisoning by antifungal antibiotics, systemically used, intentional self-harm |
| ICD10 | T36.8x2 | Poisoning by other systemic antibiotics, intentional self-harm |
| ICD10 | T36.92x | Poisoning by unspecified systemic antibiotic, intentional self-harm |
| ICD10 | T37.0x2 | Poisoning by sulfonamides, intentional self-harm |
| ICD10 | T37.1x2 | Poisoning by antimycobacterial drugs, intentional self-harm |
| ICD10 | T37.2x2 | Poisoning by antimalarials and drugs acting on other blood protozoa, intentional self-harm |
| ICD10 | T37.3x2 | Poisoning by other antiprotozoal drugs, intentional self-harm |
| ICD10 | T37.4x2 | Poisoning by anthelminthics, intentional self-harm |
| ICD10 | T37.5x2 | Poisoning by antiviral drugs, intentional self-harm |
| ICD10 | T37.8x2 | Poisoning by other specified systemic anti-infectives and antiparasitics, intentional self-harm |
| ICD10 | T37.92x | Poisoning by unspecified systemic anti-infective and antiparasitics, intentional self-harm |
| ICD10 | T38.0x2 | Poisoning by glucocorticoids and synthetic analogues, intentional self-harm |
| ICD10 | T38.1x2 | Poisoning by thyroid hormones and substitutes, intentional self-harm |
| ICD10 | T38.2x2 | Poisoning by antithyroid drugs, intentional self-harm |
| ICD10 | T38.3x2 | Poisoning by insulin and oral hypoglycemic [antidiabetic] drugs, intentional self-harm |
| ICD10 | T38.4x2 | Poisoning by oral contraceptives, intentional self-harm |
| ICD10 | T38.5x2 | Poisoning by other estrogens and progestogens, intentional self-harm |
| ICD10 | T38.6x2 | Poisoning by antigonadotrophins, antiestrogens, antiandrogens, not elsewhere classified, intentional self-harm |
| ICD10 | T38.7x2 | Poisoning by androgens and anabolic congeners, intentional self-harm |
| ICD10 | T38.802 | Poisoning by unspecified hormones and synthetic substitutes, intentional self-harm |
| ICD10 | T38.812 | Poisoning by anterior pituitary [adenohypophyseal] hormones, intentional self-harm |
| ICD10 | T38.892 | Poisoning by other hormones and synthetic substitutes, intentional self-harm |
| ICD10 | T38.902 | Poisoning by unspecified hormone antagonists, intentional self-harm |
| ICD10 | T38.992 | Poisoning by other hormone antagonists, intentional self-harm |
| ICD10 | T39.012 | Poisoning by aspirin, intentional self-harm |
| ICD10 | T39.092 | Poisoning by salicylates, intentional self-harm |
| ICD10 | T39.1x2 | Poisoning by 4-Aminophenol derivatives, intentional self-harm |
| ICD10 | T39.2x2 | Poisoning by pyrazolone derivatives, intentional self-harm |
| ICD10 | T39.312 | Poisoning by propionic acid derivatives, intentional self-harm |
| ICD10 | T39.392 | Poisoning by other nonsteroidal anti-inflammatory drugs [NSAID], intentional self-harm |
| ICD10 | T39.4x2 | Poisoning by antirheumatics, not elsewhere classified, intentional self-harm |
| ICD10 | T39.8x2 | Poisoning by other nonopioid analgesics and antipyretics, not elsewhere classified, intentional self-harm |
| ICD10 | T39.92x | Poisoning by unspecified nonopioid analgesic, antipyretic and antirheumatic, intentional self-harm |
| ICD10 | T40.0x2 | Poisoning by opium, intentional self-harm |

| | | |
|---|---|---|
| ICD10 | T40.1x2 | Poisoning by heroin, intentional self-harm |
| ICD10 | T40.2x2 | Poisoning by other opioids, intentional self-harm |
| ICD10 | T40.3x2 | Poisoning by methadone, intentional self-harm |
| ICD10 | T40.4x2 | Poisoning by other synthetic narcotics, intentional self-harm |
| ICD10 | T40.5x2 | Poisoning by cocaine, intentional self-harm |
| ICD10 | T40.602 | Poisoning by unspecified narcotics, intentional self-harm |
| ICD10 | T40.692 | Poisoning by other narcotics, intentional self-harm |
| ICD10 | T40.7x2 | Poisoning by cannabis (derivatives), intentional self-harm |
| ICD10 | T40.8x2 | Poisoning by lysergide [LSD], intentional self-harm |
| ICD10 | T40.902 | Poisoning by unspecified psychodysleptics [hallucinogens], intentional self-harm |
| ICD10 | T40.992 | Poisoning by other psychodysleptics [hallucinogens], intentional self-harm |
| ICD10 | T41.0x2 | Poisoning by inhaled anesthetics, intentional self-harm |
| ICD10 | T41.1x2 | Poisoning by intravenous anesthetics, intentional self-harm |
| ICD10 | T41.202 | Poisoning by unspecified general anesthetics, intentional self-harm |
| ICD10 | T41.292 | Poisoning by other general anesthetics, intentional self-harm |
| ICD10 | T41.3x2 | Poisoning by local anesthetics, intentional self-harm |
| ICD10 | T41.42x | Poisoning by unspecified anesthetic, intentional self-harm |
| ICD10 | T41.5x2 | Poisoning by therapeutic gases, intentional self-harm |
| ICD10 | T42.0x2 | Poisoning by hydantoin derivatives, intentional self-harm |
| ICD10 | T42.1x2 | Poisoning by iminostilbenes, intentional self-harm |
| ICD10 | T42.2x2 | Poisoning by succinimides and oxazolidinediones, intentional self-harm |
| ICD10 | T42.3x2 | Poisoning by barbiturates, intentional self-harm |
| ICD10 | T42.4x2 | Poisoning by benzodiazepines, intentional self-harm |
| ICD10 | T42.5x2 | Poisoning by mixed antiepileptics, intentional self-harm |
| ICD10 | T42.6x2 | Poisoning by other antiepileptic and sedative-hypnotic drugs, intentional self-harm |
| ICD10 | T42.72x | Poisoning by unspecified antiepileptic and sedative-hypnotic drugs, intentional self-harm |
| ICD10 | T42.8x2 | Poisoning by antiparkinsonism drugs and other central muscle-tone depressants, intentional self-harm |
| ICD10 | T43.012 | Poisoning by tricyclic antidepressants, intentional self-harm |
| ICD10 | T43.022 | Poisoning by tetracyclic antidepressants, intentional self-harm |
| ICD10 | T43.1x2 | Poisoning by monoamine-oxidase-inhibitor antidepressants, intentional self-harm |
| ICD10 | T43.202 | Poisoning by unspecified antidepressants, intentional self-harm |
| ICD10 | T43.212 | Poisoning by selective serotonin and norepinephrine reuptake inhibitors, intentional self-harm |
| ICD10 | T43.222 | Poisoning by selective serotonin reuptake inhibitors, intentional self-harm |
| ICD10 | T43.292 | Poisoning by other antidepressants, intentional self-harm |
| ICD10 | T43.3x2 | Poisoning by phenothiazine antipsychotics and neuroleptics, intentional self-harm |
| ICD10 | T43.4x2 | Poisoning by butyrophenone and thiothixene neuroleptics, intentional self-harm |
| ICD10 | T43.502 | Poisoning by unspecified antipsychotics and neuroleptics, intentional self-harm |
| ICD10 | T43.592 | Poisoning by other antipsychotics and neuroleptics, intentional self-harm |
| ICD10 | T43.602 | Poisoning by unspecified psychostimulants, intentional self-harm |
| ICD10 | T43.612 | Poisoning by caffeine, intentional self-harm |
| ICD10 | T43.622 | Poisoning by amphetamines, intentional self-harm |
| ICD10 | T43.632 | Poisoning by methylphenidate, intentional self-harm |
| ICD10 | T43.692 | Poisoning by other psychostimulants, intentional self-harm |
| ICD10 | T43.8x2 | Poisoning by other psychotropic drugs, intentional self-harm |
| ICD10 | T43.92x | Poisoning by unspecified psychotropic drug, intentional self-harm |

| | | |
|---|---|---|
| ICD10 | T44.0x2 | Poisoning by anticholinesterase agents, intentional self-harm |
| ICD10 | T44.1x2 | Poisoning by other parasympathomimetics [cholinergics], intentional self-harm |
| ICD10 | T44.2x2 | Poisoning by ganglionic blocking drugs, intentional self-harm |
| ICD10 | T44.3x2 | Poisoning by other parasympatholytics [anticholinergics and antimuscarinics] and spasmolytics, intentional self-harm |
| ICD10 | T44.4x2 | Poisoning by predominantly alpha-adrenoreceptor agonists, intentional self-harm |
| ICD10 | T44.5x2 | Poisoning by predominantly beta-adrenoreceptor agonists, intentional self-harm |
| ICD10 | T44.6x2 | Poisoning by alpha-adrenoreceptor antagonists, intentional self-harm |
| ICD10 | T44.7x2 | Poisoning by beta-adrenoreceptor antagonists, intentional self-harm |
| ICD10 | T44.8x2 | Poisoning by centrally-acting and adrenergic-neuron-blocking agents, intentional self-harm |
| ICD10 | T44.902 | Poisoning by unspecified drugs primarily affecting the autonomic nervous system, intentional self-harm |
| ICD10 | T44.992 | Poisoning by other drug primarily affecting the autonomic nervous system, intentional self-harm |
| ICD10 | T45.0x2 | Poisoning by antiallergic and antiemetic drugs, intentional self-harm |
| ICD10 | T45.1x2 | Poisoning by antineoplastic and immunosuppressive drugs, intentional self-harm |
| ICD10 | T45.2x2 | Poisoning by vitamins, intentional self-harm |
| ICD10 | T45.3x2 | Poisoning by enzymes, intentional self-harm |
| ICD10 | T45.4x2 | Poisoning by iron and its compounds, intentional self-harm |
| ICD10 | T45.512 | Poisoning by anticoagulants, intentional self-harm |
| ICD10 | T45.522 | Poisoning by antithrombotic drugs, intentional self-harm |
| ICD10 | T45.602 | Poisoning by unspecified fibrinolysis-affecting drugs, intentional self-harm |
| ICD10 | T45.612 | Poisoning by thrombolytic drug, intentional self-harm |
| ICD10 | T45.622 | Poisoning by hemostatic drug, intentional self-harm |
| ICD10 | T45.692 | Poisoning by other fibrinolysis-affecting drugs, intentional self-harm |
| ICD10 | T45.7x2 | Poisoning by anticoagulant antagonists, vitamin K and other coagulants, intentional self-harm |
| ICD10 | T45.8x2 | Poisoning by other primarily systemic and hematological agents, intentional self-harm |
| ICD10 | T45.92x | Poisoning by unspecified primarily systemic and hematological agent, intentional self-harm |
| ICD10 | T46.0x2 | Poisoning by cardiac-stimulant glycosides and drugs of similar action, intentional self-harm |
| ICD10 | T46.1x2 | Poisoning by calcium-channel blockers, intentional self-harm |
| ICD10 | T46.2x2 | Poisoning by other antidysrhythmic drugs, intentional self-harm |
| ICD10 | T46.3x2 | Poisoning by coronary vasodilators, intentional self-harm |
| ICD10 | T46.4x2 | Poisoning by angiotensin-converting-enzyme inhibitors, intentional self-harm |
| ICD10 | T46.5x2 | Poisoning by other antihypertensive drugs, intentional self-harm |
| ICD10 | T46.6x2 | Poisoning by antihyperlipidemic and antiarteriosclerotic drugs, intentional self-harm |
| ICD10 | T46.7x2 | Poisoning by peripheral vasodilators, intentional self-harm |
| ICD10 | T46.8x2 | Poisoning by antivaricose drugs, including sclerosing agents, intentional self-harm |
| ICD10 | T46.902 | Poisoning by unspecified agents primarily affecting the cardiovascular system, intentional self-harm |
| ICD10 | T46.992 | Poisoning by other agents primarily affecting the cardiovascular system, intentional self-harm |
| ICD10 | T47.0x2 | Poisoning by histamine H2-receptor blockers, intentional self-harm |
| ICD10 | T47.1x2 | Poisoning by other antacids and anti-gastric-secretion drugs, intentional self-harm |
| ICD10 | T47.2x2 | Poisoning by stimulant laxatives, intentional self-harm |

| | | |
|---|---|---|
| ICD10 | T47.3x2 | Poisoning by saline and osmotic laxatives, intentional self-harm |
| ICD10 | T47.4x2 | Poisoning by other laxatives, intentional self-harm |
| ICD10 | T47.5x2 | Poisoning by digestants, intentional self-harm |
| ICD10 | T47.6x2 | Poisoning by antidiarrheal drugs, intentional self-harm |
| ICD10 | T47.7x2 | Poisoning by emetics, intentional self-harm |
| ICD10 | T47.8x2 | Poisoning by other agents primarily affecting gastrointestinal system, intentional self-harm |
| ICD10 | T47.92x | Poisoning by unspecified agents primarily affecting the gastrointestinal system, intentional self-harm |
| ICD10 | T48.0x2 | Poisoning by oxytocic drugs, intentional self-harm |
| ICD10 | T48.1x2 | Poisoning by skeletal muscle relaxants [neuromuscular blocking agents], intentional self-harm |
| ICD10 | T48.202 | Poisoning by unspecified drugs acting on muscles, intentional self-harm |
| ICD10 | T48.292 | Poisoning by other drugs acting on muscles, intentional self-harm |
| ICD10 | T48.3x2 | Poisoning by antitussives, intentional self-harm |
| ICD10 | T48.4x2 | Poisoning by expectorants, intentional self-harm |
| ICD10 | T48.5x2 | Poisoning by other anti-common-cold drugs, intentional self-harm |
| ICD10 | T48.6x2 | Poisoning by antiasthmatics, intentional self-harm |
| ICD10 | T48.902 | Poisoning by unspecified agents primarily acting on the respiratory system, intentional self-harm |
| ICD10 | T48.992 | Poisoning by other agents primarily acting on the respiratory system, intentional self-harm |
| ICD10 | T49.0x2 | Poisoning by local antifungal, anti-infective and anti-inflammatory drugs, intentional self-harm |
| ICD10 | T49.1x2 | Poisoning by antipruritics, intentional self-harm |
| ICD10 | T49.2x2 | Poisoning by local astringents and local detergents, intentional self-harm |
| ICD10 | T49.3x2 | Poisoning by emollients, demulcents and protectants, intentional self-harm |
| ICD10 | T49.4x2 | Poisoning by keratolytics, keratoplastics, and other hair treatment drugs and preparations, intentional self-harm |
| ICD10 | T49.5x2 | Poisoning by ophthalmological drugs and preparations, intentional self-harm |
| ICD10 | T49.6x2 | Poisoning by otorhinolaryngological drugs and preparations, intentional self-harm |
| ICD10 | T49.7x2 | Poisoning by dental drugs, topically applied, intentional self-harm |
| ICD10 | T49.8x2 | Poisoning by other topical agents, intentional self-harm |
| ICD10 | T49.92x | Poisoning by unspecified topical agent, intentional self-harm |
| ICD10 | T50.0x2 | Poisoning by mineralocorticoids and their antagonists, intentional self-harm |
| ICD10 | T50.1x2 | Poisoning by loop [high-ceiling] diuretics, intentional self-harm |
| ICD10 | T50.2x2 | Poisoning by carbonic-anhydrase inhibitors, benzothiadiazides and other diuretics, intentional self-harm |
| ICD10 | T50.3x2 | Poisoning by electrolytic, caloric and water-balance agents, intentional self-harm |
| ICD10 | T50.4x2 | Poisoning by drugs affecting uric acid metabolism, intentional self-harm |
| ICD10 | T50.5x2 | Poisoning by appetite depressants, intentional self-harm |
| ICD10 | T50.6x2 | Poisoning by antidotes and chelating agents, intentional self-harm |
| ICD10 | T50.7x2 | Poisoning by analeptics and opioid receptor antagonists, intentional self-harm |
| ICD10 | T50.8x2 | Poisoning by diagnostic agents, intentional self-harm |
| ICD10 | T50.902 | Poisoning by unspecified drugs, medicaments and biological substances, intentional self-harm |
| ICD10 | T50.992 | Poisoning by other drugs, medicaments and biological substances, intentional self-harm |

| | | |
|---|---|---|
| ICD10 | T50.A12 | Poisoning by pertussis vaccine, including combinations with a pertussis component, intentional self-harm |
| ICD10 | T50.A22 | Poisoning by mixed bacterial vaccines without a pertussis component, intentional self-harm |
| ICD10 | T50.A92 | Poisoning by other bacterial vaccines, intentional self-harm |
| ICD10 | T50.B12 | Poisoning by smallpox vaccines, intentional self-harm |
| ICD10 | T50.B92 | Poisoning by other viral vaccines, intentional self-harm |
| ICD10 | T50.Z12 | Poisoning by immunoglobulin, intentional self-harm |
| ICD10 | T50.Z92 | Poisoning by other vaccines and biological substances, intentional self-harm |
| ICD10 | T51.0x2 | Toxic effect of ethanol, intentional self-harm |
| ICD10 | T51.1x2 | Toxic effect of methanol, intentional self-harm |
| ICD10 | T51.2x2 | Toxic effect of 2-Propanol, intentional self-harm |
| ICD10 | T51.3x2 | Toxic effect of fusel oil, intentional self-harm |
| ICD10 | T51.8x2 | Toxic effect of other alcohols, intentional self-harm |
| ICD10 | T51.92x | Toxic effect of unspecified alcohol, intentional self-harm |
| ICD10 | T52.0x2 | Toxic effect of petroleum products, intentional self-harm |
| ICD10 | T52.1x2 | Toxic effect of benzene, intentional self-harm |
| ICD10 | T52.2x2 | Toxic effect of homologues of benzene, intentional self-harm |
| ICD10 | T52.3x2 | Toxic effect of glycols, intentional self-harm |
| ICD10 | T52.4x2 | Toxic effect of ketones, intentional self-harm |
| ICD10 | T52.8x2 | Toxic effect of other organic solvents, intentional self-harm |
| ICD10 | T52.92x | Toxic effect of unspecified organic solvent, intentional self-harm |
| ICD10 | T53.0x2 | Toxic effect of carbon tetrachloride, intentional self-harm |
| ICD10 | T53.1x2 | Toxic effect of chloroform, intentional self-harm |
| ICD10 | T53.2x2 | Toxic effect of trichloroethylene, intentional self-harm |
| ICD10 | T53.3x2 | Toxic effect of tetrachloroethylene, intentional self-harm |
| ICD10 | T53.4x2 | Toxic effect of dichloromethane, intentional self-harm |
| ICD10 | T53.5x2 | Toxic effect of chlorofluorocarbons, intentional self-harm |
| ICD10 | T53.6x2 | Toxic effect of other halogen derivatives of aliphatic hydrocarbons, intentional self-harm |
| ICD10 | T53.7x2 | Toxic effect of other halogen derivatives of aromatic hydrocarbons, intentional self-harm |
| ICD10 | T53.92x | Toxic effect of unspecified halogen derivatives of aliphatic and aromatic hydrocarbons, intentional self-harm |
| ICD10 | T54.0x2 | Toxic effect of phenol and phenol homologues, intentional self-harm |
| ICD10 | T54.1x2 | Toxic effect of other corrosive organic compounds, intentional self-harm |
| ICD10 | T54.2x2 | Toxic effect of corrosive acids and acid-like substances, intentional self-harm |
| ICD10 | T54.3x2 | Toxic effect of corrosive alkalis and alkali-like substances, intentional self-harm |
| ICD10 | T54.92x | Toxic effect of unspecified corrosive substance, intentional self-harm |
| ICD10 | T55.0x2 | Toxic effect of soaps, intentional self-harm |
| ICD10 | T55.1x2 | Toxic effect of detergents, intentional self-harm |
| ICD10 | T56.0x2 | Toxic effect of lead and its compounds, intentional self-harm |
| ICD10 | T56.1x2 | Toxic effect of mercury and its compounds, intentional self-harm |
| ICD10 | T56.2x2 | Toxic effect of chromium and its compounds, intentional self-harm |
| ICD10 | T56.3x2 | Toxic effect of cadmium and its compounds, intentional self-harm |
| ICD10 | T56.4x2 | Toxic effect of copper and its compounds, intentional self-harm |
| ICD10 | T56.5x2 | Toxic effect of zinc and its compounds, intentional self-harm |
| ICD10 | T56.6x2 | Toxic effect of tin and its compounds, intentional self-harm |
| ICD10 | T56.7x2 | Toxic effect of beryllium and its compounds, intentional self-harm |

| | | |
|---|---|---|
| ICD10 | T56.812 | Toxic effect of thallium, intentional self-harm |
| ICD10 | T56.892 | Toxic effect of other metals, intentional self-harm |
| ICD10 | T56.92x | Toxic effect of unspecified metal, intentional self-harm |
| ICD10 | T57.0x2 | Toxic effect of arsenic and its compounds, intentional self-harm |
| ICD10 | T57.1x2 | Toxic effect of phosphorus and its compounds, intentional self-harm |
| ICD10 | T57.2x2 | Toxic effect of manganese and its compounds, intentional self-harm |
| ICD10 | T57.3x2 | Toxic effect of hydrogen cyanide, intentional self-harm |
| ICD10 | T57.8x2 | Toxic effect of other specified inorganic substances, intentional self-harm |
| ICD10 | T57.92x | Toxic effect of unspecified inorganic substance, intentional self-harm |
| ICD10 | T58.02x | Toxic effect of carbon monoxide from motor vehicle exhaust, intentional self-harm |
| ICD10 | T58.12x | Toxic effect of carbon monoxide from utility gas, intentional self-harm |
| ICD10 | T58.2x2 | Toxic effect of carbon monoxide from incomplete combustion of other domestic fuels, intentional self-harm |
| ICD10 | T58.8x2 | Toxic effect of carbon monoxide from other source, intentional self-harm |
| ICD10 | T58.92x | Toxic effect of carbon monoxide from unspecified source, intentional self-harm |
| ICD10 | T59.0x2 | Toxic effect of nitrogen oxides, intentional self-harm |
| ICD10 | T59.1x2 | Toxic effect of sulfur dioxide, intentional self-harm |
| ICD10 | T59.2x2 | Toxic effect of formaldehyde, intentional self-harm |
| ICD10 | T59.3x2 | Toxic effect of lacrimogenic gas, intentional self-harm |
| ICD10 | T59.4x2 | Toxic effect of chlorine gas, intentional self-harm |
| ICD10 | T59.5x2 | Toxic effect of fluorine gas and hydrogen fluoride, intentional self-harm |
| ICD10 | T59.6x2 | Toxic effect of hydrogen sulfide, intentional self-harm |
| ICD10 | T59.7x2 | Toxic effect of carbon dioxide, intentional self-harm |
| ICD10 | T59.812 | Toxic effect of smoke, intentional self-harm |
| ICD10 | T59.892 | Toxic effect of other specified gases, fumes and vapors, intentional self-harm |
| ICD10 | T59.92x | Toxic effect of unspecified gases, fumes and vapors, intentional self-harm |
| ICD10 | T60.0x2 | Toxic effect of organophosphate and carbamate insecticides, intentional self-harm |
| ICD10 | T60.1x2 | Toxic effect of halogenated insecticides, intentional self-harm |
| ICD10 | T60.2x2 | Toxic effect of other insecticides, intentional self-harm |
| ICD10 | T60.3x2 | Toxic effect of herbicides and fungicides, intentional self-harm |
| ICD10 | T60.4x2 | Toxic effect of rodenticides, intentional self-harm |
| ICD10 | T60.8x2 | Toxic effect of other pesticides, intentional self-harm |
| ICD10 | T60.92x | Toxic effect of unspecified pesticide, intentional self-harm |
| ICD10 | T61.02x | Ciguatera fish poisoning, intentional self-harm |
| ICD10 | T61.12x | Scombroid fish poisoning, intentional self-harm |
| ICD10 | T61.772 | Other fish poisoning, intentional self-harm |
| ICD10 | T61.782 | Other shellfish poisoning, intentional self-harm |
| ICD10 | T61.8x2 | Toxic effect of other seafood, intentional self-harm |
| ICD10 | T61.92x | Toxic effect of unspecified seafood, intentional self-harm |
| ICD10 | T62.0x2 | Toxic effect of ingested mushrooms, intentional self-harm |
| ICD10 | T62.1x2 | Toxic effect of ingested berries, intentional self-harm |
| ICD10 | T62.2x2 | Toxic effect of other ingested (parts of) plant(s), intentional self-harm |
| ICD10 | T62.8x2 | Toxic effect of other specified noxious substances eaten as food, intentional self-harm |
| ICD10 | T62.92x | Toxic effect of unspecified noxious substance eaten as food, intentional self-harm |
| ICD10 | T63.002 | Toxic effect of unspecified snake venom, intentional self-harm |
| ICD10 | T63.012 | Toxic effect of rattlesnake venom, intentional self-harm |
| ICD10 | T63.022 | Toxic effect of coral snake venom, intentional self-harm |
| ICD10 | T63.032 | Toxic effect of taipan venom, intentional self-harm |

| | | |
|---|---|---|
| ICD10 | T63.042 | Toxic effect of cobra venom, intentional self-harm |
| ICD10 | T63.062 | Toxic effect of venom of other North and South American snake, intentional self-harm |
| ICD10 | T63.072 | Toxic effect of venom of other Australian snake, intentional self-harm |
| ICD10 | T63.082 | Toxic effect of venom of other African and Asian snake, intentional self-harm |
| ICD10 | T63.092 | Toxic effect of venom of other snake, intentional self-harm |
| ICD10 | T63.112 | Toxic effect of venom of gila monster, intentional self-harm |
| ICD10 | T63.122 | Toxic effect of venom of other venomous lizard, intentional self-harm |
| ICD10 | T63.192 | Toxic effect of venom of other reptiles, intentional self-harm |
| ICD10 | T63.2x2 | Toxic effect of venom of scorpion, intentional self-harm |
| ICD10 | T63.302 | Toxic effect of unspecified spider venom, intentional self-harm |
| ICD10 | T63.312 | Toxic effect of venom of black widow spider, intentional self-harm |
| ICD10 | T63.322 | Toxic effect of venom of tarantula, intentional self-harm |
| ICD10 | T63.332 | Toxic effect of venom of brown recluse spider, intentional self-harm |
| ICD10 | T63.392 | Toxic effect of venom of other spider, intentional self-harm |
| ICD10 | T63.412 | Toxic effect of venom of centipedes and venomous millipedes, intentional self-harm |
| ICD10 | T63.422 | Toxic effect of venom of ants, intentional self-harm |
| ICD10 | T63.432 | Toxic effect of venom of caterpillars, intentional self-harm |
| ICD10 | T63.442 | Toxic effect of venom of bees, intentional self-harm |
| ICD10 | T63.452 | Toxic effect of venom of hornets, intentional self-harm |
| ICD10 | T63.462 | Toxic effect of venom of wasps, intentional self-harm |
| ICD10 | T63.482 | Toxic effect of venom of other arthropod, intentional self-harm |
| ICD10 | T63.512 | Toxic effect of contact with stingray, intentional self-harm |
| ICD10 | T63.592 | Toxic effect of contact with other venomous fish, intentional self-harm |
| ICD10 | T63.612 | Toxic effect of contact with Portuguese Man-o-war, intentional self-harm |
| ICD10 | T63.622 | Toxic effect of contact with other jellyfish, intentional self-harm |
| ICD10 | T63.632 | Toxic effect of contact with sea anemone, intentional self-harm |
| ICD10 | T63.692 | Toxic effect of contact with other venomous marine animals, intentional self-harm |
| ICD10 | T63.712 | Toxic effect of contact with venomous marine plant, intentional self-harm |
| ICD10 | T63.792 | Toxic effect of contact with other venomous plant, intentional self-harm |
| ICD10 | T63.812 | Toxic effect of contact with venomous frog, intentional self-harm |
| ICD10 | T63.822 | Toxic effect of contact with venomous toad, intentional self-harm |
| ICD10 | T63.832 | Toxic effect of contact with other venomous amphibian, intentional self-harm |
| ICD10 | T63.892 | Toxic effect of contact with other venomous animals, intentional self-harm |
| ICD10 | T63.92x | Toxic effect of contact with unspecified venomous animal, intentional self-harm |
| ICD10 | T64.02x | Toxic effect of aflatoxin, intentional self-harm |
| ICD10 | T64.82x | Toxic effect of other mycotoxin food contaminants, intentional self-harm |
| ICD10 | T65.0x2 | Toxic effect of cyanides, intentional self-harm |
| ICD10 | T65.1x2 | Toxic effect of strychnine and its salts, intentional self-harm |
| ICD10 | T65.212 | Toxic effect of chewing tobacco, intentional self-harm |
| ICD10 | T65.222 | Toxic effect of tobacco cigarettes, intentional self-harm |
| ICD10 | T65.292 | Toxic effect of other tobacco and nicotine, intentional self-harm |
| ICD10 | T65.3x2 | Toxic effect of nitroderivatives and aminoderivatives of benzene and its homologues, intentional self-harm |
| ICD10 | T65.4x2 | Toxic effect of carbon disulfide, intentional self-harm |
| ICD10 | T65.5x2 | Toxic effect of nitroglycerin and other nitric acids and esters, intentional self-harm |
| ICD10 | T65.6x2 | Toxic effect of paints and dyes, not elsewhere classified, intentional self-harm |
| ICD10 | T65.812 | Toxic effect of latex, intentional self-harm |
| ICD10 | T65.822 | Toxic effect of harmful algae and algae toxins, intentional self-harm |

| | | |
|---|---|---|
| ICD10 | T65.832 | Toxic effect of fiberglass, intentional self-harm |
| ICD10 | T65.892 | Toxic effect of other specified substances, intentional self-harm |
| ICD10 | T65.92x | Toxic effect of unspecified substance, intentional self-harm |
| ICD10 | T71.112 | Asphyxiation due to smothering under pillow, intentional self-harm |
| ICD10 | T71.122 | Asphyxiation due to plastic bag, intentional self-harm |
| ICD10 | T71.132 | Asphyxiation due to being trapped in bed linens, intentional self-harm |
| ICD10 | T71.152 | Asphyxiation due to smothering in furniture, intentional self-harm |
| ICD10 | T71.162 | Asphyxiation due to hanging, intentional self-harm |
| ICD10 | T71.192 | Asphyxiation due to mechanical threat to breathing due to other causes, intentional self-harm |
| ICD10 | T71.222 | Asphyxiation due to being trapped in a car trunk, intentional self-harm |
| ICD10 | T71.232 | Asphyxiation due to being trapped in a (discarded) refrigerator, intentional self-harm |
| ICD10 | T14.91 | Suicide attempt |
| ICD10 | R45.851 | Suicidal ideation |

**Supplemental Information for NLP manual validation at the individual note level**. The UUHSC "cases" in this validation comprised 22 suicide deaths (11 male, 11 female) with evidence from ICD-9 or ICD-10 codes for at least one previous instance of non-lethal suicidal behaviors, including attempts and/or ideation prior to the fatal attempt. These cases were hypothesized to have positive mention of suicidality from the NLP.

The "controls" were selected from a larger pool of population controls in our ongoing studies where there were at least two encounters resulting in an ICD-9 or ICD-10 code of depression, but no ICD-9 or ICD-10 codes indicating suicide attempt or suicidal ideation. Controls were matched for sex, and were within 2 years to suicide deaths. Because of multiple depression diagnoses, controls enriched to potentially have relevant notes for the NLP, but had unknown positivity of suicidality mentions in the notes.

All cases and controls were White of non-Hispanic ethnicity. Average age was xx and xx for cases and controls, respectively.

**Supplemental Table S2. Characteristics of EHR notes in the manual validation of the NLP**. Notes were labeled according to source (psychiatric note vs. not psychiatric note) so that results could be characterized according to psychiatric vs. more general note source. Numbers and sources of notes are described in supplemental table S3.

We used the original lexicon developed for this NLP, with the exception that we removed "sa" and "si" occurring without other text. These terms created false positives in notes from non-psychiatric encounters.

| Note Characteristic | Cases | Controls | Total |
|---|---|---|---|
| Total N of notes | 6,249 | 9,099 | 15,349 |
| N of psychiatric encounter notes | 4,750 (76%) | 6,096 (67%) | 10,846 |
| N of notes from other sources | 1,500 (24%) | 3,003 (33%) | 4,503 |
| N notes manually annotated | 293 | 111 | 404 |
| N psych notes manually annotated | 222 | 53 | 275 |
| N other notes manually annotated | 71 | 58 | 129 |

**Supplemental Table S3. Validation details of the NLP within individual notes by note type (psychiatric vs. non-psychiatric encounter)**. As expected, notes for suicides with ICD diagnoses of prior non-fatal suicidality (cases) showed more true positive instances of suicidality from the NLP (60.75% for Si and 40.27% for SB) than the matched controls with depression but no ICD diagnoses of suicidality (24.32% for SI and 17.12% for SB). Notes also appeared to be more clearly either positive or negative among cases, with low instances of false positives and no instances of false negatives. Among controls, false positive and negative instances were observed, and ranged from 1% to 8%. Across both cases and controls, false positive rates were approximately 3% and false negative rates were approximately 1%.

| Validation Attribute | Cases | Controls | Total |
|---|---|---|---|
| True Positive SI, psych notes | 139/222 = 62.61% | 18/53 = 33.96% | 157/275 = 57.09% |
| False Positive SI, psych notes | 1/222 = 0.45% | 0/53 = 0 % | 1/275 = 0.36% |
| True Positive SI, non-psych notes | 39/71 = 54.92% | 9/58 = 15.12% | 48/129 = 37.21% |
| False Positive SI, non-psych notes | 4/71 = 5.63% | 6/58 = 10.34% | 10/129 = 7.75% |
| True Positive SI, all notes | 178/293 = 60.75% | 27/111 = 24.32% | 205/404 = 50.74% |
| False Positive SI, all notes | 5/293 = 1.71% | 6/111 = 5.41% | 11/404 = 2.72% |
| True Positive SB, psych notes | 89/222 = 40.10% | 15/53 = 28,30% | 104/275 = 37.82% |
| False Positive SB, psych notes | 2/222 = 0.90% | 6/53 = 11.32% | 8/275 = 2.91% |
| True Positive SB, non-psych notes | 29/71 = 40.84% | 4/58 = 6.90% | 33/129 = 25.58% |
| False Positive SB, non-psych notes | 2/71 = 2.82% | 3/58 = 5.17% | 5/129 = 3.88% |
| True Positive SB, all notes | 118/293 = 40.27% | 19/111 = 17.12% | 137/404 = 33.91% |
| False Positive SB, all notes | 4/293 = 0.68% | 9/111 = 8.12% | 13/404 = 3.22% |
| True Negative SI, psych notes | 80/222 = 36.04% | 34/53 = 64.15% | 114/275 = 41.45% |
| False Negative SI, psych notes | 0/222 = 0% | 3/53 = 5.66% | 3/275 = 1.09% |
| True Negative SI, non-psych notes | 30/71 = 42.25% | 40/58 = 68.97% | 70/129 = 54.26% |
| False Negative SI, non-psych notes | 0/71 = 0% | 3/58 = 5.17% | 3/129 = 2.33% |
| True Negative SI, all notes | 110/293 = 37.54% | 74/111 = 66.67% | 184/404 = 45.55% |

| | | | |
|---|---|---|---|
| False Negative SI, all notes | 0/293 = 0% | 6/111 = 5.41% | 6/404 = 1.49% |
| True Negative SB, psych notes | 129/222 = 58.11% | 34/53 = 64.15% | 163/275 = 59.27% |
| False Negative SB, psych notes | 0/222 = 0% | 0/53 = 0% | 0/275 = 0% |
| True Negative SB, non-psych notes | 44/71 = 61.97% | 45/58 = 77.59% | 89/129 = 68.99% |
| False Negative SB, non-psych notes | 0/71 = 0% | 1/58 = 1.72% | 1/129 = 0.78% |
| True Negative SB, all notes | 173/293 = 59.04% | 79/111 = 71.17% | 252/404 = 62.38% |
| False Negative SB, all notes | 0/293 = 0% | 1/111 = 0.90% | 1/404 = 0.25% |

**Supplemental table S4. Rater agreement details and kappa statistics**. Rater agreement was generally high, and did not differ substantially by note type (psychiatric vs. non-psychiatric).

| Rater attribute | Cases | Controls | Total |
|---|---|---|---|
| Rater agreement SI, psych notes | 178/222 = 80.18% | 50/53 = 94.34% | 228/275 = 82.91% |
| Rater agreement SI, non-psych notes | 57/71 = 80.28% | 54/58 = 93.10% | 111/129 = 86.05% |
| Rater agreement SI, all notes | 235/293 = 80.20% | 104/111 = 93.60% | 339/404 = 83.91% |
| Rater agreement SB, psych notes | 192/222 = 86.49% | 48/53 = 90.57% | 240/275 = 87.27% |
| Rater agreement SB, non-psych notes | 53/71 = 74.65% | 55/58 = 94.83% | 108/129 = 83.72% |
| Rater agreement SB, all notes | 245/293 = 83.62% | 103/111 = 92.79% | 348/404 = 86.14% |

**Supplemental table S5. Kappa statistics for rater agreement**.

| Note type | Cases | Controls | Total |
|---|---|---|---|
| SI, psych notes | 0.603 | 0.872 | 0.678 |
| SI, non-psych notes | 0.606 | 0.862 | 0.721 |
| SI, all notes | 0.604 | 0.887 | 0.745 |
| SB, psych notes | 0.730 | 0.811 | 0.745 |
| SB, non-psych notes | 0.493 | 0.897 | 0.674 |
| SB, all notes | 0.672 | 0.856 | 0.723 |

**Supplemental table S6. Final metrics from manual validation of the NLP in individual notes**.

| | SI | | | SB | | |
|---|---|---|---|---|---|---|
| Variable | Cases | Controls | All | Cases | Controls | All |
| All Note Sources | | | | | | |
| Precision[1] | 0.973 | 0.818 | 0.949 | 0.967 | 0.679 | 0.949 |
| Recall [2] | 1.0 | 0.818 | 0.972 | 1.0 | 0.950 | 0.993 |
| F1 Score[3] | 0.986 | 0.818 | 0.960 | 0.983 | 0.792 | 0.971 |
| Psychiatric Notes | | | | | | |
| Precision[1] | 0.993 | 1.0 | 0.994 | 0.978 | 0.714 | 0.929 |
| Recall [2] | 1.0 | 0.857 | 0.963 | 1.0 | 0.833 | 0.946 |
| F1 Score[3] | 0.996 | 0.923 | 0.978 | 0.989 | 0.769 | 0.937 |
| Other Non-Psych Notes | | | | | | |
| Precision[1] | 0.907 | 0.600 | 0.828 | 0.936 | 0.571 | 0.868 |
| Recall [2] | 1.0 | 0.750 | 0.972 | 1.0 | 0.571 | 0.917 |
| F1 Score[3] | 0.951 | 0.667 | 0.894 | 0.967 | 0.571 | 0.892 |

Note. Precision= positive predictive value = (true positive)/(true positive + false positive). Recall = sensitivity = true positive/(true positive + false negative). F1 score = 2*(precision*recall)/(precision+recall).

**Supplemental information for NLP validation aggregating all text for each person rather than considering individual notes**. Because our objective for the NLP is to determine suicidality at the person level we revisited our manual validation and determined rates of false positives and negatives and rater agreement when considering the entire corpus of notes for each person as the source text.

**Supplemental table S7. NLP performance considering the entire corpus of notes for each person as the source text, rather than individual notes**.

| | **SI** | | | **SB** | | |
|---|---|---|---|---|---|---|
| **Variable** | **Cases** | **Controls** | **All** | **Cases** | **Controls** | **All** |
| True positives | 20/22 = 90.91% | 7/22=31.82% | 27/44=61.36% | 21/22=95.46% | 7/22=31.82% | 28/44=63.64% |
| False positives | 0 | 1/22=4.55% | 1/44=2.27% | 0 | 2/22=9.09% | 2/44=4.55% |
| True negatives | 2/22 = 9.09% | 14/22=63.64% | 16/22=36.36% | 1/22=4.55% | 13/22=59.09% | 14/44=31.82% |
| Precision | 1.0 | 0.875 | 0.964 | 1.0 | 0.778 | 0.933 |
| Recall | 1.0 | 1.0 | 1.0 | 1.0 | 1.0 | 1.0 |
| F1 Score | 1.0 | 0.933 | 0.982 | 1.0 | 0.875 | 0.966 |

Note. There were no false negatives at the aggregated person level for either cases or controls, so this attribute is not included in the table. Precision= positive predictive value = (true positive)/(true positive + false positive). Recall = sensitivity = true positive/(true positive + false negative). F1 score = 2*(precision*recall)/(precision+recall).

Using aggregated note text, 21 of the 22 suicide deaths were true positive for SB. The one case that was not positive for SB was positive for SI. All of the suicide deaths that were identified by the NLP as positive for SB were also identified as positive for SI except two cases which were only positive for SB. Therefore, all 22 suicide deaths were positive when considering either SI or SB.

In the control records, though none had diagnostic billing codes for SB or SI, nine were found to be positive by the NLP for SB. Eight controls were also positive for SI. Manual review indicated two of the positive controls for SB were false positives, and one of the positive controls for SI was a false positive. These false positives occurred in notes where suicide terms in the lexicon rare or complex, or were negated in complex or non-standard phrasing.

Of note, defining suicidality as either SB or SI, the true positive rate was 29/44 = 65.91%, the false positive rate was 3/44 = 6.82%, and the false negative rate was 0. Precision for this overall definition was therefore 0.936 and the F1 score was 0.967.

**Supplemental Table S8. Mental health PheCodes eliminated for this study.**

| PheCode | Reason for Elimination |
|---|---|
| Suicidal ideation or attempt | High overlap with group definition codes |
| Suicide or self-Inflicted injury | High overlap with group definition codes |
| Other mental disorder | Non-specific, difficult to interpret |
| Other conditions of brain NOS | Non-specific, difficult to interpret |
| Other signs and symptoms Involving mental state | Non-specific, difficult to interpret |
| Symptoms involving head and neck | Non-specific, difficult to interpret |
| Other persistent mental disorders due to conditions classified elsewhere | Non-specific, difficult to interpret |
| Other specified nonpsychotic and/or transient mental disorders | Non-specific, difficult to interpret |
| Transient mental disorders due to conditions classified elsewhere | Non-specific, difficult to interpret |

**Supplemental Table S9. 32 Diagnostic PheCodes associated with mental health**. This table includes PheCodes with significant effects of either suicide death or of presence of SI/SB comparing suicide (SUI) to age-/birth year-matched controls (CTL).

| PheCode | ICD-9 codes | ICD-10 codes |
|---|---|---|
| Depression | 296.21, 296.31, 311 | F32.0, F32.9, F33.0 |
| Bipolar | 296, 296.01, 296.02, 296.03, 296.04, 296.05, 296.06, 296.1, 296.11, 296.12, 296.13, 296.14, 296.15, 296.16, 296.4, 296.41, 296.42, 296.43, 296.44, 296.45, 296.46, 296.5, 296.51, 296.52, 296.53, 296.54, 296.55, 296.56, 296.6, 296.61, 296.62, 296.63, 296.64, 296.65, 296.66, 296.7, 296.8, 296.81, 296.82, 296.89 | F30.1, F30.10, F30.11, F30.12, F30.13, F30.2, F30.3, F30.4, F30.8, F30.9, F31, F31.0, F31.1, F31.10, F31.11, F31.12, F31.13, F31.2, F31.3, F31.30, F31.31, F31.32, F31.4, F31.5, F31.6, F31.60, F31.61, F31.62, F31.63, F31.64, F31.7, F31.70, F31.71, F31.72, F31.73, F31.74, F31.75, F31.76, F31.77, F31.78, F31.8, F31.81, F31.89, F31.9, F32.8, F32.81, F32.89 |
| Major Depressive Disorder | 296.2, 296.22, 296.23, 296.24, 296.25, 296.26, 296.3, 296.32, 296.33, 296.34, 296.35, 296.36 | F32, F32.0, F32.1, F32.2, F32.3, F32.4, F32.5, F32.8, F32.81, F32.89, F32.9, F33, F33.0, F33.1, F33.2, F33.3, F33.4, F33.40, F33.41, F33.42, F33.8, F33.9 |
| Dysthymic disorders | 300.4 | F34.1 |
| Other mood disorders | 293.83, 296, 296.9, 296.99, V11.1 | F06.3, F06.30, F06.31, F06.32, F06.33, F06.34, F30-F39, F33.8, F34.8, F34.81, F34.89, F34.9, F39 |
| Somatoform disorder | 300.11, 300.7, 300.8, 300.81, 300.82, 307.8, 307.89 | F44.4, F44.5, F44.6, F44.7, F45, F45.0, F45.1, F45.2, F45.20, F45.21, F45.22, F45.29, F45.4, F45.41, F45.42 |
| Anxiety-related states & conditions | 300, 300.5, 300.89, 300.9, 313.1, 313.21, 313.22, 313.3, 313.82, 313.83 | F48.8, F48.9, F50-F59, F99, R45.2, R45.5, R45.6 |
| Posttraumatic stress disorder | 309.81, V11.4 | F43.1, F43.10, F43.11, F43.12, Z86.51 |
| Anxiety disorder | 293.84, 300, 300.09, 300.1, 313 | F06.4, F41.3, F41.8, F41.9 |
| Agoraphobia, social phobia, panic disorder | 300.01, 300.21, 300.22, 300.23 | F40.0, F40.00, F40.01, F40.02, F40.1, F40.10, F40.11, F41.0 |
| Obsessive-compulsive disorders | 300.3 | F42, F42.2, F42.3, F42.4, F42.8, F42.9 |
| Generalized anxiety disorder | 300.2, 300.02 | F41.1 |
| Eating disorders | 307.5, 307.51, 307.52, 307.53, 307.54, 307.59 | F50, F50.2, F50.8, F50.81, F50.82, F50.89, F50.9, F98.2, F98.21, F98.29, F98.3, G43.A |
| Acute reaction to stress | 308, 308.1, 308.2, 308.3, 308.4, 308.9 | F43.0, R45.7 |
| Adjustment reaction | 309, 309.1, 309.2, 309.22, 309.23, 309.24, 309.28, 309.29, 309.3, 309.4, 309.8. 309.82, 309.83, 309.89, 309.9 | F43.2, F43.20, F43.21, F43.22, F43.23, F43.24, F43.25, F43.29, F43.8, F43.9, F94.8 |
| Personality disorders (excluding ASP & BD) | 301, 301.1, 301.11, 301.12, 301.13, 301.5, 301.51, 301.59, 301.6, 301.8, 301.81, 301.82, 301.89, 301.9 | F34.0 F60.0, F60.4, F60.6, F60.7, F60.8, F60.81, F60.89, F60.9, F68.1, F68.10, F68.11, F68.12, F68.13, F69 |
| Antisocial & borderline PD | 301.3, 301.7, 301.83, 301.84 | F60.2, F60.3, F60.8 |

| | | |
|---|---|---|
| Conduct disorders | 312, 312.01, 312.02, 312.03, 312.1, 312.11, 312.12, 312.13, 312.2, 312.21, 312.22, 312.23, 312.4, 312.8, 312.81, 312.82, 312.89, 312.9, 313.81 | F91, F91.0, F91.1, F91.2, F91.3, F91.8, F91.9, Y09 |
| Attention deficit hyperactivity disorder | 314, 314.01, 314.1, 314.2, 314.8, 314.9 | F90, F90.0, F90.1, F90.2, F90.8, F90.9 |
| Developmental delays & disorders | 307.9, 315, 315.4, 315.5, 315.8, 315.9, V40 | F01-F99, F63.3, F81.9, F82, F88, R45.1, R45.81, R45.82 |
| Delirium | 291, 292.81, 293, 293.1 | F05, F10.121, F10.221, F10.231, F10.921, F11.121, F11.221, F11.921, F12.121, F12.221, F12.921, F13.121, F13.221, F13.921, F14.121, F14.221, F14.921, F15.121, F15.221, F15.921, F16.121, F16.221, F16.921, F18.121, F18.221, F18.921, F19.121, F19.221, F19.921 |
| Alteration of consciousness | 780, 780.09 | R40.0, R40.1 |
| Paranoid disorders | 297, 297.1, 297.2, 297.3, 297.8, 297.9, 298.3, 298.4 | F22, F23, F24 |
| Psychosis | 293.81, 293.82, 298, 298.1, 298.8, 298.9 | F06.0, F06.2, F28, F29 |
| Schizophrenia | 295, 295.01, 295.02, 295.03, 295.04, 295.05, 295.1, 295.11, 295.12, 295.13, 295.14, 295.15, 295.2, 295.21, 295.22, 295.23, 295.24, 295.25, 295.3, 295.31, 295.32, 295.33, 295.34, 295.35, 295.4, 295.41, 295.42, 295.43, 295.44, 295.45, 295.5, 295.51, 295.52, 295.53, 295.54, 295.55, 295.6, 295.61, 295.62, 295.63, 295.64, 295.65, 295.7, 295.71, 295.72, 295.73, 295.74, 295.75, 295.8, 295.81, 295.82, 295.83, 295.84, 295.85, 295.9, 295.91, 295.92, 295.93, 295.94, 295.95, V11.0 | F20, F20.0, F20.1, F20.2, F20.3, F20.5, F20.8, F20.81, F20.89, F20.9, F25, F25.0, F25.1, F25.8, F25.9 |
| Altered mental status | 780.97 | R41.0, R41.82 |
| Hallucinations | 780.1 | R44.0, R44.2, R44.3 |
| Substance addiction & disorders | 292, 292.1, 292.11, 292.12, 292.2, 292.8, 292.82, 292.83, 292.84, 292.85, 292.89, 292.9, 304, 304.01, 304.02, 304.03, 304.1, 304.11, 304.12, 304.13, 304.2, 304.21, 304.22, 304.23, 304.3, 304.31, 304.32, 304.33, 304.4, 304.41, 304.42, 304.43, 304.5, 304.51, 304.52, 304.53, 304.6, 304.61, 304.62, 304.63, 304.7, 304.71, 304.72, 304.73, 304.8, 304.81, 304.82, 304.83, 304.9, 304.91, 304.92, 304.93, 305, 305.2, 305.21, 305.22, 305.23, 305.3, 305.31, 305.32, 305.33, 305.4, 305.41, 305.42, 305.43, 305.5, 305.51, 305.52, 305.53, 305.6, 305.61, 305.62, 305.63, 305.7, 305.71, 305.72, 305.73, 305.8, 305.81, 305.82, 305.83, 305.9, 305.91, 305.92, 305.93, 306, 648.3, 648.31, 648.32, 648.33, 648.34, 965, 965.01, 965.02 | F11, F11.1, F11.10, F11.120, F11.122, F11.129, F11.14, F11.150, F11.151, F11.159, F11.181, F11.182, F11.188, F11.19, F11.2, F11.20, F11.21, F11.220, F11.221, F11.222, F11.229, F11.23, F11.24, F11.250, F11.251, F11.259, F11.281, F11.282, F11.288, F11.29, F11.90, F11.920, F11.922, F11.929, F11.93, F11.94, F11.950, F11.951, F11.959, F11.981, F11.982, F11.988, F11.99, F12, F12.1, F12.10, F12.120, F12.122, F12.129, F12.150, F12.151, F12.159, F12.180, F12.188, F12.19, F12.2, F12.20, F12.21, F12.220, F12.221, F12.222, F12.229, F12.250, F12.251, F12.259, F12.280, F12.288, F12.29, F12.90, F12.920, F12.922, F12.929, F12.950, F12.951, F12.959, F12.980, F12.988, F12.99, F13, F13.10, F13.120, F13.129, F13.14, F13.150, F13.151, F13.159, F13.180, F13.181, |

| | | |
|---|---|---|
| | | F13.182, F13.188, F13.19, F13.20, F13.21, F13.220, F13.221, F13.229, F13.230, F13.231, F13.232, F13.239, F13.24, F13.250, F13.251, F13.259, F13.26, F13.27, F13.280, F13.281, F13.282, F13.288, F13.29, F13.90, F13.920, F13.929, F13.930, F13.931, F13.932, F13.939, F13.94, F13.950, F13.951, F13.959, F13.96, F13.97, F13.980, F13.981, F13.982, F13.988, F13.99, F14, F14.1, F14.10, F14.120, F14.122, F14.129, F14.14, F14.150, F14.151, F14.159, F14.180, F14.181, F14.182, F14.188, F14.19, F14.2, F14.20, F14.21, F14.220, F14.221, F14.222, F14.229, F14.23, F14.24, F14.250, F14.251, F14.259, F14.280, F14.281, F14.282, F14.288, F14.29, F14.90, F14.920, F14.922, F14.929, F14.94, F14.950, F14.951, F14.959, F14.980, F14.981, F14.982, F14.988, F14.99, F15, F15.10, F15.120, F15.122, F15.129, F15.14, F15.150, F15.151, F15.159, F15.180, F15.181, F15.182, F15.188, F15.19, F15.20, F15.21, F15.220, F15.221, F15.222, F15.229, F15.23, F15.24, F15.250, F15.251, F15.259, F15.280, F15.281, F15.282, F15.288, F15.29, F15.90, F15.920, F15.922, F15.929, F15.93, F15.94, F15.950, F15.951, F15.959, F15.980, F15.981, F15.982, F15.988, F15.99, F16, F16.1, F16.10, F16.120, F16.122, F16.129, F16.14, F16.150, F16.151, F16.159, F16.180, F16.183, F16.188, F16.19, F16.2, F16.20, F16.21, F16.220, F16.221, F16.229, F16.24, F16.250, F16.251, F16.259, F16.280, F16.283, F16.288, F16.29, F16.90, F16.920, F16.929, F16.94, F16.950, F16.951, F16.959, F16.980, F16.983, F16.988, F16.99, F17.203, F17.208, F17.209, F17.213, F17.218, F17.219, F17.223, F17.228, F17.229, F17.293, F17.298, F17.299, F18, F18.1, F18.10, F18.120, F18.129, F18.14, F18.150, F18.151, F18.159, F18.17, F18.180, F18.188, F18.19, F18.2, F18.20, F18.21, F18.220, F18.221, F18.229, F18.24, F18.250, F18.251, F18.259, F18.27, F18.280, F18.288, F18.29, F18.90, F18.920, F18.929, F18.94, F18.950, F18.951, F18.959, F18.97, F18.980, F18.988, F18.99, F19.10, F19.120, F19.122, F19.129, F19.14, F19.150, F19.151, F19.159, F19.16, F19.17, F19.180, F19.181, F19.182, F19.188, F19.19, F19.20, F19.21, F19.220, F19.221, F19.222, F19.229, F19.230, F19.231, F19.232, F19.239, F19.24, F19.250, F19.251, F19.259, F19.26, F19.27, F19.280, F19.281, F19.282, F19.288, F19.29, F19.90, F19.920, F19.922, F19.929, F19.930, F19.931, F19.932, F19.939, F19.94, F19.950, F19.951, F19.959, F19.96, F19.97, |

| | | |
|---|---|---|
| | | F19.980, F19.981, F19.982, F19.988, F19.99, F55, F55.0, F55.1, F55.2, F55.3, F55.4, F55.8, O99.32, O99.320, O99.321, O99.322, O99.323, O99.324, O99.325, T40.0X1, T40.0X1A, T40.0X2, T40.0X2A, T40.0X3, T40.0X3A, T40.0X4, T40.0X4A, T40.1X1, T40.1X1, T40.1X1A, T40.1X2, T40.1X2A, T40.1X3, T40.1X3A, T40.1X4, T40.1X4A, T40.3X1, T40.3X1, T40.3X1A, T40.3X2, T40.3X2A, T40.3X3, T40.3X3A, T40.3X4, T40.3X4A |
| Alcoholism | 291.1, 291.2, 291.3, 291.81, 305, 305.01, 305.02, 305.03, 305.5, 375.5, 535.3, 535.31 | F10.1, F10.10, F10.120, F10.129, F10.151, F10.230, F10.232, F10.239, F10.251, F10.26, F10.27, F10.951, F10.96, F10.97, G62.1, K29.2, K29.20, K29.21, V11.3, Z65, Z65.0, Z65.1, Z65.2, Z65.3, Z65.4, Z65.5, Z65.8, Z65.9 |
| Alcohol-related disorders | 291, 291.4, 291.5, 291.8, 291.82, 291.89, 291.9, 303, 303.01, 303.02, 303.03, 303.9, 303.91, 303.92, 303.93, 790.3, 980, E860.0, E860.1 | F10, F10.14, F10.150, F10.159, F10.180, F10.181, F10.182, F10.188, F10.19, F10.2, F10.20, F10.21, F10.22, F10.220, F10.229, F10.24, F10.250, F10.259, F10.280, F10.281, F10.282, F10.288, F10.29, F10.920, F10.929, F10.94, F10.950, F10.959, F10.980, F10.981, F10.982, F10.988, F10.99, R78.0, T51.0, T51.0X, T51.0X1, T51.0X1, T51.0X1A, T51.0X2, T51.0X2A, T51.0X3, T51.0X3A, T51.0X4, T51.0X4A |
| Adverse events of opiates & narcotics in therapeutic use | 965, 965.09, E850.0, E850.1, E850.2, E935.0, E935.1, E935.2, V14.5 | T40.0X1, T40.0X5, T40.0X5A, T40.0X5D, T40.0X5S, T40.1X1, T40.1X1A, T40.1X5A, T40.1X5S, T40.2X1, T40.2X1A, T40.2X2, T40.2X2A, T40.2X3, T40.2X3A, T40.2X4, T40.2X4A, T40.2X5, T40.2X5A, T40.2X5D, T40.2X5S, T40.3X1, T40.3X1A, T40.3X5, T40.3X5A, T40.3X5D, T40.3X5S, T40.4X1, T40.4X1A, T40.4X2, T40.4X2A, T40.4X3, T40.4X3A, T40.4X4, T40.4X4A, T40.4X5, T40.4X5A, T40.4X5D, T40.4X5S, T40.5X5, T40.5X5A, T40.5X5D, T40.5X5S, T40.601, T40.601A, T40.602, T40.602A, T40.603, T40.603A, T40.604, T40.604A, T40.605, T40.605A, T40.605D, T40.605S, T40.691, T40.691A, T40.692, T40.692A, T40.693, T40.693A, T40.694, T40.694A, T40.695, T40.695A, T40.695D, T40.695S, T40.7X5, T40.7X5A, T40.7X5D, T40.7X5S, T40.905, T40.905A, T40.905D, T40.905S, T40.995, T40.995A, T40.995D, T40.995S, Z88.5 |
| Tobacco use disorder | 305.1, 305.11, 305.12, 305.13, 649, 649.01, 649.02, 649.03, 649.04, V15.82 | F17.200, F17.201, F17.202, F17.210, F17.211, F17.220, F17.221, F17.290, F17.291, O99.33, O99.330, O99.331, O99.332, O99.333, O99.334, O99.335, Z72.0, Z87.891 |

Supplemental Figure 1. Prevalences of mental health PheCodes adjusted for age, sex, adjusted overall N of PheCodes: a) affective disorders; b) anxiety/stress disorders; c) personality, impulsivity, developmental disorders; d) psychotic disorders; e) substance use disorders.

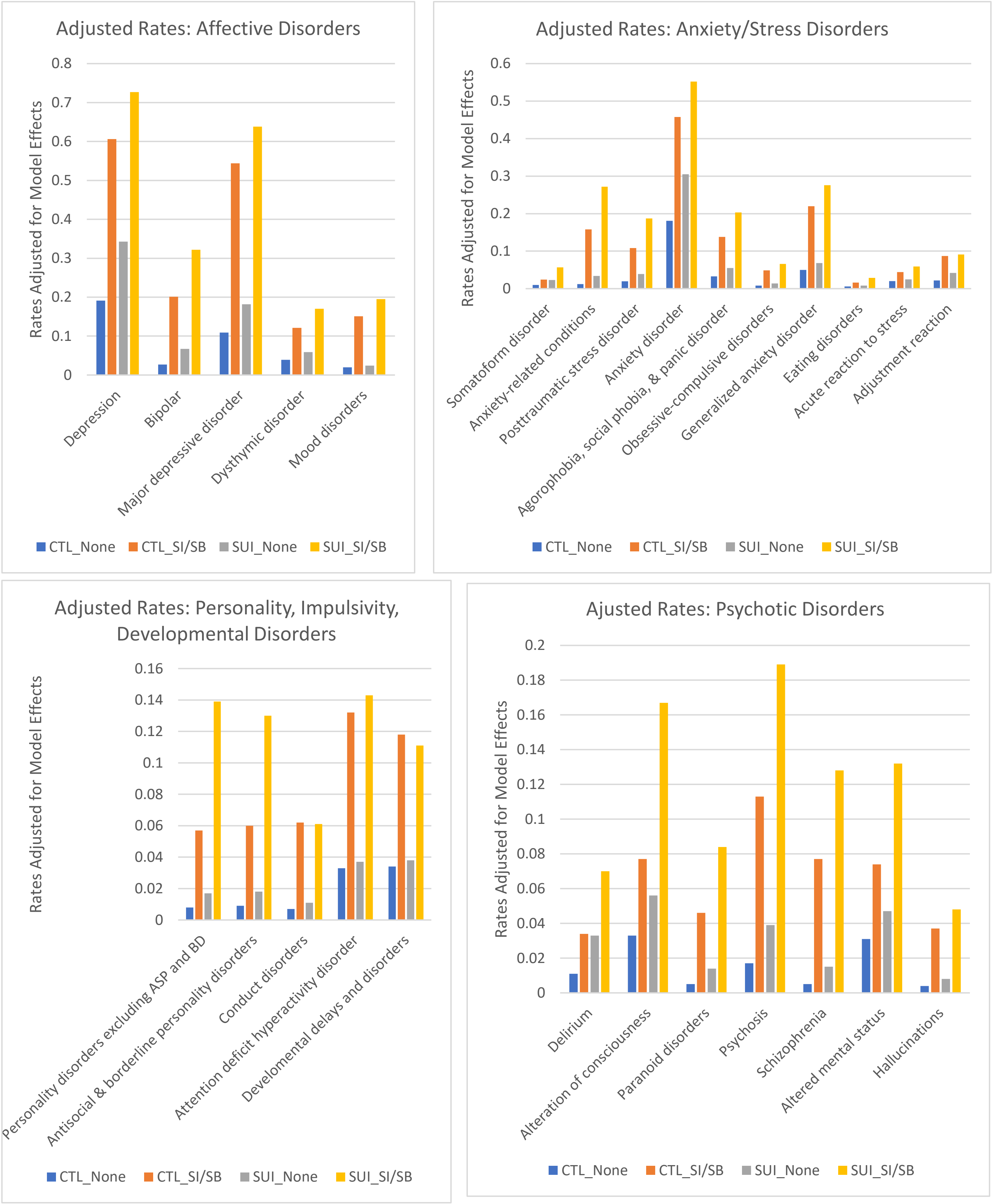

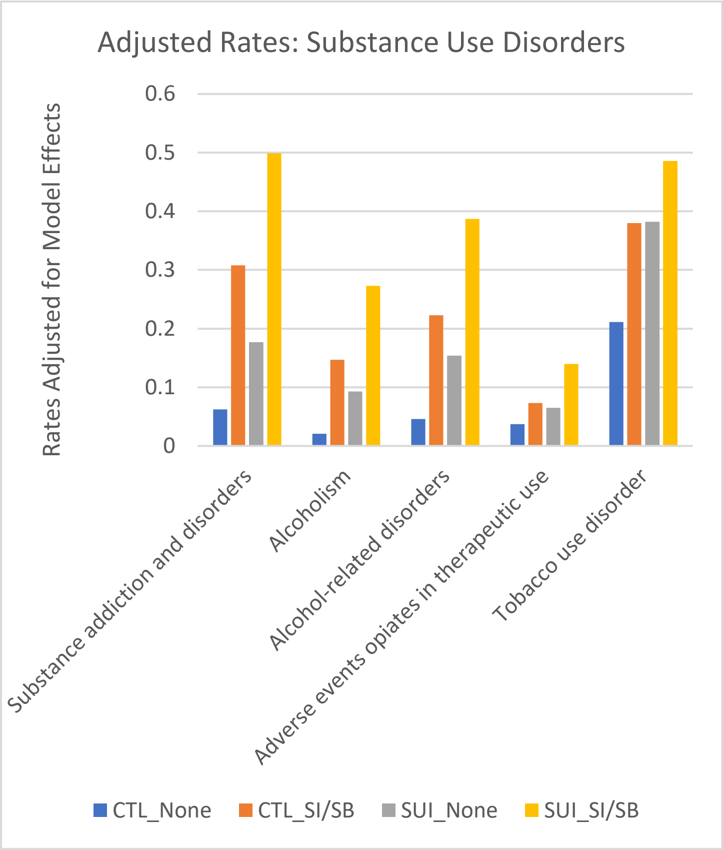
Adjusted Rates: Substance Use Disorders
Rates Adjusted for Model Effects
0.6
0.5
0.4
0.3
0.2
0.1
0
Substance addiction and disorders
Alcoholism
Alcohol-related disorders
Adverse events opiates in therapeutic use
Tobacco use disorder
CTL_None
CTL_SI/SB
SUI_None
SUI_SI/SB

**Table S10. Standardized marginal effects of logistic regressions of physical health PheCodes where there were negative (protective) effects of having SI/SB or of suicide death.**

| PheCode | Grouping | Effect: suicide death | p-value | Effect: presence SI/SB | p-value |
|---|---|---|---|---|---|
| Atrial fibrillation | Circulatory | 0.156 | 1.04E-01 | **-0.448** | **4.52E-06** |
| Encounter for long-term (current) use of anticoagulants | Circulatory | 0.129 | 1.12E-01 | **-0.529** | **1.35E-11** |
| Coronary atherosclerosis | Circulatory | 0.086 | 2.81E-01 | **-0.457** | **9.87E-09** |
| Other venous embolism and thrombosis | Circulatory | 0.076 | 3.67E-01 | **-0.327** | **3.73E-05** |
| Hemorrhoids | Circulatory | -0.078 | 1.65E-01 | **-0.229** | **1.31E-05** |
| Encounter for long-term (current) use of aspirin | Circulatory | -0.170 | 3.85E-02 | **-0.357** | **4.45E-06** |
| Cardiomegaly | Circulatory | -0.246 | 8.66E-03 | **-0.534** | **4.85E-10** |
| Rash and other nonspecific skin eruption | Dermatologic | **-0.284** | **8.35E-06** | -0.180 | 1.08E-03 |
| Atopic/contact dermatitis due to other or unspecified | Dermatologic | **-0.330** | **4.03E-08** | **-0.286** | **4.58E-08** |
| Actinic keratosis | Dermatologic | **-0.384** | **6.07E-06** | **-0.712** | **2.17E-18** |
| Acne | Dermatologic | **-0.442** | **1.83E-08** | -0.047 | 4.65E-01 |
| Sebaceous cyst | Dermatologic | **-0.473** | **1.33E-06** | **-0.417** | **5.36E-07** |
| Seborrheic keratosis | Dermatologic | **-0.608** | **7.13E-13** | **-0.740** | **3.49E-22** |
| Other dyschromia | Dermatologic | **-0.907** | **1.13E-16** | **-0.537** | **5.58E-10** |
| Other diseases of the teeth and supporting structures | Digestive | 0.069 | 4.37E-01 | **0.453** | **2.15E-08** |
| Diverticulosis | Digestive | -0.093 | 1.81E-01 | **-0.331** | **9.66E-07** |
| Dysphagia | Digestive | -0.096 | 1.81E-01 | **-0.293** | **6.34E-06** |
| Bariatric surgery | Digestive | **-0.421** | **1.76E-09** | **-0.312** | **2.29E-07** |
| Hyperlipidemia | Endocrine | **-0.244** | **1.76E-05** | **-0.294** | **3.32E-08** |
| Morbid obesity | Endocrine | **-0.313** | **1.29E-05** | -0.074 | 2.38E-01 |
| Overweight, obesity and other hyperalimentation | Endocrine | **-0.353** | **6.72E-06** | 0.209 | 2.11E-03 |
| Obesity | Endocrine | **-0.452** | **1.78E-14** | -0.033 | 5.10E-01 |
| Other disorders of the kidney and ureters | Genitourinary | 0.054 | 4.85E-01 | **-0.301** | **3.84E-05** |
| Other anemias | Hematopoietic | 0.143 | 2.03E-02 | **-0.393** | **5.30E-12** |
| Thrombocytopenia | Hematopoietic | 0.087 | 3.72E-01 | **-0.370** | **3.77E-05** |
| Lymphadenitis | Hematopoietic | -0.225 | 2.33E-03 | **-0.335** | **1.96E-07** |
| Septicemia | Infectious | 0.115 | 2.15E-01 | **-0.441** | **2.26E-07** |
| Fever of unknown origin | Infectious | -0.120 | 2.53E-02 | **-0.461** | **2.16E-22** |
| Bacterial infection NOS | Infectious | -0.159 | 1.18E-01 | **-0.445** | **5.46E-07** |
| Conjunctivitis, infectious | Infectious | **-0.431** | **7.69E-09** | **-0.274** | **1.16E-05** |
| Influenza | Infectious | **-0.443** | **4.90E-07** | -0.264 | 2.74E-04 |
| Viral warts & HPV | Infectious | **-0.447** | **6.78E-08** | **-0.446** | **1.03E-10** |
| Sprains and strains | Injuries | 0.013 | 7.63E-01 | **-0.175** | **1.25E-05** |
| Dislocation | Injuries | -0.053 | 3.87E-01 | **-0.398** | **1.01E-12** |
| Internal derangement of knee | Injuries | -0.136 | 5.70E-02 | **-0.415** | **1.91E-10** |
| Joint/ligament sprain | Injuries | -0.149 | 3.29E-02 | **-0.259** | **2.12E-05** |
| Fracture of unspecified bones | Injuries | -0.158 | 1.36E-02 | **-0.457** | **6.79E-16** |
| Other derangement of joint | Injuries | -0.174 | 2.86E-02 | **-0.412** | **4.01E-09** |
| Allergies, other | Injuries | **-0.372** | **3.40E-05** | 0.111 | 1.46E-01 |
| Arthropathy NOS | Musculoskeletal | 0.087 | 2.14E-01 | **-0.269** | **5.75E-05** |
| Osteopenia or other disorder of bone and cartilage | Musculoskeletal | 0.043 | 5.85E-01 | **-0.469** | **1.60E-10** |
| Osteoarthrosis NOS | Musculoskeletal | 0.004 | 9.51E-01 | **-0.266** | **6.66E-06** |

| | | | | | |
|---|---|---|---|---|---|
| Osteoarthritis; localized | Musculoskeletal | -0.008 | 9.20E-01 | **-0.433** | **2.18E-09** |
| Joint effusions | Musculoskeletal | -0.034 | 6.19E-01 | **-0.457** | **1.71E-13** |
| Peripheral enthesopathies and allied syndromes | Musculoskeletal | -0.110 | 9.71E-02 | **-0.324** | **1.26E-07** |
| Symptoms and disorders of the joints | Musculoskeletal | -0.118 | 1.91E-01 | **-0.568** | **2.02E-12** |
| Synovitis and tenosynovitis | Musculoskeletal | -0.147 | 6.51E-02 | **-0.388** | **8.30E-08** |
| Pain in joint | Musculoskeletal | **-0.187** | **1.38E-05** | **-0.355** | **1.99E-19** |
| Pain in limb | Musculoskeletal | **-0.215** | **2.97E-06** | **-0.198** | **1.52E-06** |
| Other disorders of bone and cartilage | Musculoskeletal | -0.279 | 1.67E-03 | **-0.440** | **1.96E-08** |
| Osteoarthrosis, localized, primary | Musculoskeletal | **-0.337** | **1.12E-05** | **-0.376** | **8.43E-08** |
| Enthesopathy | Musculoskeletal | **-0.446** | **1.71E-08** | **-0.509** | **8.97E-14** |
| Benign neoplasm of colon | Neoplasms | **-0.322** | **2.45E-07** | **-0.352** | **1.77E-09** |
| Neoplasm of uncertain behavior of skin | Neoplasms | **-0.367** | **8.17E-05** | **-0.546** | **6.58E-11** |
| Acquired absence of breast | Neoplasms | **-0.401** | **1.88E-05** | -0.202 | 1.21E-02 |
| Neoplasm of uncertain behavior | Neoplasms | **-0.510** | **1.28E-09** | **-0.573** | **1.96E-15** |
| Benign neoplasm of skin | Neoplasms | **-0.605** | **1.15E-22** | **-0.626** | **6.53E-32** |
| Radiotherapy | Neoplasms | **-0.675** | **2.32E-13** | **-0.404** | **9.06E-08** |
| Screening for malignant neoplasms of the skin | Neoplasms | **-0.885** | **3.86E-43** | **-0.233** | **1.45E-05** |
| Chemotherapy | Neoplasms | **-0.913** | **3.28E-28** | **-0.504** | **2.32E-14** |
| Sleep disorders | Neurologic | -0.173 | 3.63E-02 | **0.351** | **1.52E-06** |
| Obstructive sleep apnea | Neurologic | **-0.287** | **1.74E-05** | -0.074 | 2.15E-01 |
| Obstetrical/birth trauma | Pregnancy conditions | **-0.500** | **2.12E-07** | **-0.496** | **7.97E-10** |
| Other conditions or status of the mother complicating pregnancy, childbirth, or the puerperium | Pregnancy conditions | **-0.604** | **3.08E-09** | **-0.533** | **4.04E-10** |
| Other complications of pregnancy NEC | Pregnancy conditions | **-0.692** | **4.38E-08** | **-0.493** | **1.72E-06** |
| Acute bronchitis and bronchiolitis | Respiratory | -0.154 | 6.82E-03 | **-0.204** | **4.52E-05** |
| Other upper respiratory disease | Respiratory | -0.181 | 2.40E-03 | **-0.257** | **9.60E-07** |
| Acute sinusitis | Respiratory | **-0.308** | **1.80E-08** | **-0.363** | **3.21E-14** |
| Allergic rhinitis | Respiratory | **-0.359** | **3.09E-08** | **-0.313** | **1.96E-08** |
| Acute upper respiratory infections of multiple or unspecified sites | Respiratory | **-0.398** | **3.42E-17** | **-0.290** | **1.72E-12** |
| Cough | Respiratory | **-0.446** | **1.31E-19** | **-0.241** | **2.06E-08** |
| Acute pharyngitis | Respiratory | **-0.448** | **7.17E-21** | **-0.201** | **1.01E-06** |
| Suppurative and unspecified otitis media | Sense organs | -0.230 | 5.00E-04 | **-0.377** | **2.34E-11** |
| Presbyopia | Sense organs | **-0.423** | **1.92E-06** | -0.127 | 1.11E-01 |
| Astigmatism | Sense organs | **-0.594** | **1.46E-15** | -0.174 | 5.01E-03 |
| Myopia | Sense organs | **-0.723** | **1.30E-23** | **-0.272** | **3.47E-06** |
| Thoracic or lumbosacral neuritis or radiculitis, unspecified | Symptoms | 0.182 | 1.04E-02 | **-0.275** | **3.89E-05** |
| Musculoskeletal symptoms referable to limbs | Symptoms | 0.039 | 6.63E-01 | **-0.418** | **4.68E-07** |
| Abdominal pain | Symptoms | 0.026 | 5.53E-01 | **-0.174** | **9.83E-06** |
| Malaise and fatigue | Symptoms | -0.009 | 8.46E-01 | **-0.180** | **2.91E-05** |
| Edema | Symptoms | -0.028 | 6.74E-01 | **-0.293** | **2.06E-06** |
| Muscle weakness | Symptoms | -0.097 | 3.60E-01 | **-0.473** | **6.92E-07** |

Notes. Standardized marginal effects were derived from logistic regressions.[37] Each binary presence/absence PheCode was regressed on case-control status (suicide death effect) and then on presence/absence of SI/SB (presence SI/SB effect), adjusting for age, sex, and transformed overall number of PheCodes. Tests that were significant after adjustment for multiple testing are shown in bold type.

**Table S11. Differences in prevalences for mental health PheCodes when testing the parsimonious model with covariates of age, sex, and overall number of diagnoses (transformed) vs. testing the model including additional covariates of age*age, age*sex, and time period cohort[1].**

| | | Adjusted prevalences with age, sex, N codes | | | | Adjusted prevalences with age, sex, N codes, age*age, age*sex, time period cohort | | | | Difference in adjusted prevalences (additional covariate model – original model) | | | |
|---|---|---|---|---|---|---|---|---|---|---|---|---|---|
| PheCode | Grouping | CTL_ None | CTL_SI/ SB | SUI_ None | SUI_SI/ SB | CTL_ None | CTL_SI/ SB | SUI_ None | SUI_SI/ SB | diff CTL_ None | diff CTL_SI/ SB | diff SUI_ None | diff SUI_SI /SB |
| Depression | affective | 0.191 | 0.606 | 0.343 | 0.726 | 0.201 | 0.614 | 0.348 | 0.724 | 0.01 | 0.008 | 0.005 | -0.002 |
| Bipolar | affective | 0.027 | 0.201 | 0.067 | 0.322 | 0.038 | 0.209 | 0.072 | 0.324 | 0.011 | 0.008 | 0.005 | 0.002 |
| Major depressive disorder | affective | 0.109 | 0.545 | 0.182 | 0.638 | 0.101 | 0.535 | 0.179 | 0.639 | -0.008 | -0.01 | -0.003 | 0.001 |
| Dysthymic disorder | affective | 0.039 | 0.121 | 0.059 | 0.17 | 0.051 | 0.131 | 0.065 | 0.172 | 0.012 | 0.01 | 0.006 | 0.002 |
| Mood disorders | affective | 0.019 | 0.151 | 0.024 | 0.195 | 0.02 | 0.152 | 0.024 | 0.195 | 0.001 | 0.001 | 0 | 0 |
| Somatoform disorder | anx/stress | 0.01 | 0.024 | 0.023 | 0.057 | 0.017 | 0.031 | 0.027 | 0.059 | 0.007 | 0.007 | 0.004 | 0.002 |
| Anxiety-related conditions | anx/stress | 0.012 | 0.158 | 0.034 | 0.272 | 0.029 | 0.173 | 0.043 | 0.278 | 0.017 | 0.015 | 0.009 | 0.006 |
| Posttraumatic stress disorder | anx/stress | 0.02 | 0.109 | 0.039 | 0.187 | 0.022 | 0.111 | 0.041 | 0.188 | 0.0025 | 0.002 | 0.002 | 0.001 |
| Anxiety disorder | anx/stress | 0.181 | 0.458 | 0.305 | 0.551 | 0.173 | 0.449 | 0.3 | 0.542 | -0.008 | -0.009 | -0.005 | -0.009 |
| Agorophobia, social phobia, & panic disorder | anx/stress | 0.033 | 0.138 | 0.055 | 0.203 | 0.037 | 0.141 | 0.057 | 0.203 | 0.004 | 0.003 | 0.002 | 0 |
| Obsessive-compulsive disorders | anx/stress | 0.008 | 0.049 | 0.014 | 0.066 | 0.009 | 0.051 | 0.015 | 0.067 | 0.001 | 0.002 | 0.001 | 0.001 |
| Generalized anxiety disorder | anx/stress | 0.051 | 0.22 | 0.068 | 0.276 | 0.041 | 0.21 | 0.063 | 0.273 | -0.01 | -0.01 | -0.005 | -0.003 |
| Eating disorders | anx/stress | 0.006 | 0.016 | 0.008 | 0.029 | 0.007 | 0.017 | 0.008 | 0.029 | 0.001 | 0.001 | 0 | 0 |
| Acute reaction to stress | anx/stress | 0.02 | 0.044 | 0.025 | 0.059 | 0.024 | 0.048 | 0.027 | 0.06 | 0.004 | 0.004 | 0.002 | 0.001 |
| Adjustment reaction | anx/stress | 0.023 | 0.089 | 0.042 | 0.091 | 0.028 | 0.094 | 0.046 | 0.095 | 0.005 | 0.005 | 0.004 | 0.004 |
| Personality disorders excluding ASP and BD | PD | 0.008 | 0.059 | 0.017 | 0.141 | 0.019 | 0.069 | 0.023 | 0.145 | 0.011 | 0.01 | 0.006 | 0.004 |
| Antisocial & borderline personality disorders | PD | 0.009 | 0.06 | 0.018 | 0.13 | 0.019 | 0.069 | 0.024 | 0.134 | 0.01 | 0.009 | 0.006 | 0.004 |
| Conduct disorders | impulsive | 0.007 | 0.061 | 0.011 | 0.061 | 0.007 | 0.062 | 0.011 | 0.062 | 0 | 0.001 | 0 | 0.001 |

| | | | | | | | | | | | | | |
|---|---|---|---|---|---|---|---|---|---|---|---|---|---|
| Attention deficit hyperactivity disorder | impulsive | 0.033 | 0.132 | 0.037 | 0.143 | 0.025 | 0.125 | 0.033 | 0.142 | -0.008 | -0.007 | -0.004 | -0.001 |
| Develomental delays and disorders | developmental | 0.034 | 0.118 | 0.038 | 0.111 | 0.028 | 0.112 | 0.037 | 0.112 | -0.006 | -0.006 | -0.001 | 0.001 |
| Delirium | psychotic | 0.011 | 0.034 | 0.033 | 0.07 | 0.021 | 0.043 | 0.039 | 0.074 | 0.01 | 0.009 | 0.006 | 0.004 |
| Alteration of consciousness | psychotic | 0.033 | 0.076 | 0.056 | 0.167 | 0.05 | 0.092 | 0.065 | 0.172 | 0.017 | 0.016 | 0.009 | 0.005 |
| Paranoid disorders | psychotic | 0.005 | 0.046 | 0.014 | 0.084 | 0.006 | 0.046 | 0.015 | 0.084 | 0.001 | 0 | 0.001 | 0 |
| Psychosis | psychotic | 0.018 | 0.113 | 0.039 | 0.189 | 0.03 | 0.124 | 0.046 | 0.194 | 0.012 | 0.011 | 0.007 | 0.005 |
| Schizophrenia | psychotic | 0.005 | 0.077 | 0.015 | 0.128 | 0.013 | 0.084 | 0.02 | 0.132 | 0.008 | 0.007 | 0.005 | 0.004 |
| Altered mental status | psychotic | 0.031 | 0.075 | 0.047 | 0.132 | 0.033 | 0.077 | 0.048 | 0.133 | 0.002 | 0.002 | 0.001 | 0.001 |
| Hallucinations | psychotic | 0.004 | 0.037 | 0.008 | 0.048 | 0.006 | 0.039 | 0.01 | 0.049 | 0.002 | 0.002 | 0.002 | 0.001 |
| Substance addiction and disorders | sud | 0.062 | 0.309 | 0.177 | 0.499 | 0.086 | 0.328 | 0.19 | 0.505 | 0.024 | 0.019 | 0.013 | 0.006 |
| Alcoholism | sud | 0.021 | 0.148 | 0.093 | 0.273 | 0.033 | 0.157 | 0.1 | 0.276 | 0.012 | 0.009 | 0.007 | 0.003 |
| Alcohol-related disorders | sud | 0.047 | 0.223 | 0.154 | 0.387 | 0.056 | 0.23 | 0.159 | 0.386 | 0.009 | 0.007 | 0.005 | -0.001 |
| Adverse events opiates in therapeutic use | sud | 0.037 | 0.074 | 0.065 | 0.14 | 0.05 | 0.085 | 0.072 | 0.144 | 0.013 | 0.011 | 0.007 | 0.004 |
| Tobacco use disorder | sud | 0.212 | 0.38 | 0.382 | 0.486 | 0.208 | 0.375 | 0.377 | 0.472 | -0.004 | -0.005 | -0.005 | -0.014 |

[1] The cohort effect was defined in 5-year increments, with the first grouping including extra years of 1996 and 1997 because of sparse electronic health records data from those years (1996-2002, 2003-2007, 2008-2012, 2013-2017, 2018-2022).

**Table S12. Differences in prevalences for physical health PheCodes when testing the parsimonious model with covariates of age, sex, and overall number of diagnoses (transformed) vs. testing the model including additional covariates of age*age, age*sex, and time period cohort.**[1]

| | | Adjusted prevalences with age, sex, N codes | | | | Adjusted prevalences with age, sex, N codes, age*age, age*sex, time period cohort | | | | Difference in adjusted prevalences (additional covariate model – original model) | | | |
|---|---|---|---|---|---|---|---|---|---|---|---|---|---|
| PheCode | Grouping | CTL_ None | CTL_SI/ SB | SUI_ None | SUI_SI/ SB | CTL_ None | CTL_SI/ SB | SUI_ None | SUI_SI/ SB | diff CTL_ None | diff CTL_SI/ SB | diff SUI_ None | diff SUI_SI/ SB |
| Tachycardia NOS | Circulatory | 0.078 | 0.098 | 0.084 | 0.138 | 0.085 | 0.105 | 0.089 | 0.142 | 0.007 | 0.007 | 0.005 | 0.004 |
| Acute pancreatitis | Digestive | 0.021 | 0.025 | 0.038 | 0.043 | 0.027 | 0.03 | 0.042 | 0.045 | 0.006 | 0.005 | 0.004 | 0.002 |
| Peptic ulcer | Digestive | 0.033 | 0.038 | 0.049 | 0.071 | 0.049 | 0.052 | 0.057 | 0.077 | 0.016 | 0.014 | 0.008 | 0.006 |
| Hematemesis | Digestive | 0.012 | 0.018 | 0.016 | 0.035 | 0.016 | 0.022 | 0.018 | 0.037 | 0.004 | 0.004 | 0.002 | 0.002 |
| Esophagitis, GERD, and related diseases | Digestive | 0.054 | 0.039 | 0.064 | 0.062 | 0.061 | 0.045 | 0.068 | 0.063 | 0.007 | 0.006 | 0.004 | 0.001 |
| Diseases of teeth, supporting structures | Digestive | 0.027 | 0.056 | 0.036 | 0.049 | 0.031 | 0.059 | 0.038 | 0.049 | 0.004 | 0.003 | 0.002 | 0 |
| Acidosis | Endocrine | 0.037 | 0.047 | 0.047 | 0.107 | 0.046 | 0.055 | 0.052 | 0.111 | 0.009 | 0.008 | 0.005 | 0.004 |
| Hypopotassemia | Endocrine | 0.076 | 0.097 | 0.102 | 0.174 | 0.091 | 0.11 | 0.111 | 0.18 | 0.015 | 0.013 | 0.009 | 0.006 |
| Protein-calorie malnutrition | Endocrine | 0.03 | 0.04 | 0.046 | 0.058 | 0.044 | 0.053 | 0.054 | 0.067 | 0.014 | 0.013 | 0.008 | 0.009 |
| Hypovolemia | Endocrine | 0.138 | 0.157 | 0.192 | 0.199 | 0.16 | 0.178 | 0.205 | 0.211 | 0.022 | 0.021 | 0.013 | 0.012 |
| Acute renal failure | Genitourinary | 0.048 | 0.043 | 0.065 | 0.072 | 0.057 | 0.052 | 0.07 | 0.078 | 0.009 | 0.009 | 0.005 | 0.006 |
| Renal dialysis | Genitourinary | 0.014 | 0.032 | 0.016 | 0.046 | 0.017 | 0.034 | 0.019 | 0.049 | 0.003 | 0.002 | 0.003 | 0.003 |
| Urinary tract infection | Genitourinary | 0.195 | 0.174 | 0.215 | 0.211 | 0.209 | 0.188 | 0.222 | 0.219 | 0.014 | 0.014 | 0.007 | 0.008 |
| Viral hepatitis C | Infectious | 0.011 | 0.037 | 0.021 | 0.042 | 0.019 | 0.043 | 0.025 | 0.043 | 0.008 | 0.006 | 0.004 | 0.001 |
| Crushing or internal injury to organs | Injuries | 0.022 | 0.018 | 0.042 | 0.042 | 0.03 | 0.025 | 0.046 | 0.045 | 0.008 | 0.007 | 0.004 | 0.003 |
| Adverse drug events or drug allergies | Injuries | 0.059 | 0.108 | 0.069 | 0.2 | 0.075 | 0.121 | 0.078 | 0.208 | 0.016 | 0.013 | 0.009 | 0.008 |
| Fracture of ribs | Injuries | 0.028 | 0.028 | 0.047 | 0.048 | 0.034 | 0.034 | 0.05 | 0.05 | 0.006 | 0.006 | 0.003 | 0.002 |
| Fracture vertebral column w/o spinal cord injury | Injuries | 0.032 | 0.026 | 0.05 | 0.049 | 0.043 | 0.037 | 0.056 | 0.054 | 0.011 | 0.011 | 0.006 | 0.005 |
| Open wounds of extremities | Injuries | 0.077 | 0.119 | 0.11 | 0.189 | 0.084 | 0.125 | 0.114 | 0.191 | 0.007 | 0.006 | 0.004 | 0.002 |
| Concussion | Injuries | 0.049 | 0.057 | 0.082 | 0.08 | 0.049 | 0.057 | 0.082 | 0.08 | 0 | 0 | 0 | 0 |

| | | | | | | | | | | | | | |
|---|---|---|---|---|---|---|---|---|---|---|---|---|---|
| Complication of internal orthopedic device | Injuries | 0.039 | 0.024 | 0.059 | 0.034 | 0.049 | 0.033 | 0.065 | 0.038 | 0.01 | 0.009 | 0.006 | 0.004 |
| Sprains and strains of back and neck | Injuries | 0.164 | 0.206 | 0.24 | 0.237 | 0.183 | 0.22 | 0.249 | 0.236 | 0.019 | 0.014 | 0.009 | -0.001 |
| Open wound of hand | Injuries | 0.045 | 0.05 | 0.066 | 0.071 | 0.045 | 0.049 | 0.066 | 0.069 | 0 | -0.001 | 0 | -0.002 |
| Burns | Injuries | 0.034 | 0.037 | 0.051 | 0.051 | 0.039 | 0.041 | 0.054 | 0.052 | 0.005 | 0.004 | 0.003 | 0.001 |
| Superficial injury w/o infection | Injuries | 0.125 | 0.161 | 0.167 | 0.206 | 0.127 | 0.162 | 0.168 | 0.205 | 0.002 | 0.001 | 0.001 | -0.001 |
| Contusion | Injuries | 0.26 | 0.299 | 0.346 | 0.325 | 0.277 | 0.314 | 0.356 | 0.331 | 0.017 | 0.015 | 0.01 | 0.006 |
| Other open wound of head and face | Injuries | 0.027 | 0.027 | 0.032 | 0.049 | 0.034 | 0.033 | 0.036 | 0.052 | 0.007 | 0.006 | 0.004 | 0.003 |
| Skull/face fracture, intercranial injury | Injuries | 0.055 | 0.08 | 0.072 | 0.101 | 0.068 | 0.09 | 0.079 | 0.107 | 0.013 | 0.01 | 0.007 | 0.006 |
| Personal history of medication allergy | Injuries | 0.026 | 0.05 | 0.026 | 0.066 | 0.033 | 0.056 | 0.03 | 0.07 | 0.007 | 0.006 | 0.004 | 0.004 |
| Injury, NOS | Injuries | 0.248 | 0.271 | 0.292 | 0.297 | 0.249 | 0.271 | 0.293 | 0.297 | 0.001 | 0 | 0.001 | 0 |
| Other and unspecified disc disorder | Musculoskeletal | 0.039 | 0.029 | 0.064 | 0.043 | 0.049 | 0.037 | 0.069 | 0.046 | 0.01 | 0.008 | 0.005 | 0.003 |
| Degeneration of intervertebral disc | Musculoskeletal | 0.138 | 0.116 | 0.186 | 0.123 | 0.147 | 0.123 | 0.19 | 0.121 | 0.009 | 0.007 | 0.004 | -0.002 |
| Spondylosis w/o myelopathy | Musculoskeletal | 0.117 | 0.092 | 0.156 | 0.094 | 0.126 | 0.099 | 0.16 | 0.094 | 0.009 | 0.007 | 0.004 | 0 |
| Displacement of intervertebral disc | Musculoskeletal | 0.094 | 0.078 | 0.126 | 0.086 | 0.103 | 0.084 | 0.13 | 0.084 | 0.009 | 0.006 | 0.004 | -0.002 |
| Chronic pain syndrome | Neurologic | 0.015 | 0.027 | 0.029 | 0.053 | 0.019 | 0.03 | 0.031 | 0.053 | 0.004 | 0.003 | 0.002 | 0 |
| Convulsions | Neurologic | 0.049 | 0.078 | 0.083 | 0.123 | 0.073 | 0.098 | 0.097 | 0.134 | 0.024 | 0.02 | 0.014 | 0.011 |
| Encephalopathy | Neurologic | 0.021 | 0.034 | 0.025 | 0.06 | 0.027 | 0.039 | 0.029 | 0.064 | 0.006 | 0.005 | 0.004 | 0.004 |
| Unspecified disorders of nervous system | Neurologic | 0.016 | 0.025 | 0.017 | 0.051 | 0.022 | 0.03 | 0.02 | 0.054 | 0.006 | 0.005 | 0.003 | 0.003 |
| Coma | Neurologic | 0.011 | 0.028 | 0.015 | 0.045 | 0.014 | 0.029 | 0.017 | 0.047 | 0.003 | 0.001 | 0.002 | 0.002 |
| Insomnia | Neurologic | 0.108 | 0.242 | 0.155 | 0.282 | 0.103 | 0.235 | 0.153 | 0.278 | -0.005 | -0.007 | -0.002 | -0.004 |
| Chronic pain conditions | Neurologic | 0.113 | 0.169 | 0.157 | 0.195 | 0.105 | 0.16 | 0.153 | 0.19 | -0.008 | -0.009 | -0.004 | -0.005 |
| Abnormal involuntary movements | Neurologic | 0.038 | 0.06 | 0.054 | 0.078 | 0.045 | 0.066 | 0.058 | 0.081 | 0.007 | 0.006 | 0.004 | 0.003 |
| Migraine | Neurologic | 0.1 | 0.128 | 0.129 | 0.154 | 0.111 | 0.136 | 0.135 | 0.155 | 0.011 | 0.008 | 0.006 | 0.001 |

| | | | | | | | | | | | | | |
|---|---|---|---|---|---|---|---|---|---|---|---|---|---|
| Other sleep disorders | Neurologic | 0.041 | 0.067 | 0.037 | 0.047 | 0.048 | 0.072 | 0.041 | 0.05 | 0.007 | 0.005 | 0.004 | 0.003 |
| Pneumonitis due to inhalation of food or vomit | Respiratory | 0.014 | 0.027 | 0.028 | 0.086 | 0.026 | 0.037 | 0.034 | 0.091 | 0.012 | 0.01 | 0.006 | 0.005 |
| Respiratory failure | Respiratory | 0.039 | 0.041 | 0.058 | 0.108 | 0.049 | 0.05 | 0.064 | 0.113 | 0.01 | 0.009 | 0.006 | 0.005 |
| Chronic airway obstruction | Respiratory | 0.032 | 0.034 | 0.065 | 0.035 | 0.043 | 0.046 | 0.071 | 0.041 | 0.011 | 0.012 | 0.006 | 0.006 |
| Asphyxia and hypoxemia | Respiratory | 0.07 | 0.062 | 0.082 | 0.085 | 0.083 | 0.075 | 0.089 | 0.091 | 0.013 | 0.013 | 0.007 | 0.006 |
| Rhabdomyolysis | Symptoms | 0.005 | 0.015 | 0.011 | 0.047 | 0.007 | 0.017 | 0.012 | 0.047 | 0.002 | 0.002 | 0.001 | 0 |
| Syncope and collapse | Symptoms | 0.092 | 0.106 | 0.12 | 0.124 | 0.101 | 0.115 | 0.125 | 0.128 | 0.009 | 0.009 | 0.005 | 0.004 |
| Cervicalgia | Symptoms | 0.144 | 0.145 | 0.178 | 0.169 | 0.154 | 0.152 | 0.183 | 0.169 | 0.01 | 0.007 | 0.005 | 0 |

[1] The cohort effect was defined in 5-year increments, with the first grouping including extra years of 1996 and 1997 because of sparse electronic health records data from those years (1996-2002, 2003-2007, 2008-2012, 2013-2017, 2018-2022).

**Supplementary Table S13. Characteristics of prevalences of suicide deaths for mental health PheCodes where there was a significant effect of violent method of death vs. non-violent method of death.**

| PheCode | Grouping | SUI_None_ non-violent | Sui_SI/SB_ non-violent | SUI_ None_violent | SUI_SI/SB_ violent | p-value (violent effect) |
|---|---|---|---|---|---|---|
| Major depressive disorder | Affective | 0.428 | 0.789 | 0.366 | 0.754 | 0.0005 |
| Anxiety related conditions | Anx/stress | 0.077 | 0.305 | 0.055 | 0.244 | 0.001 |
| Substance addiction and disorders | SUD | 0.290 | 0.534 | 0.225 | 0.483 | 0.0001 |
| Adverse events of opioids in therapeutic use | SUD | 0.142 | 0.164 | 0.095 | 0.127 | <0.0001 |
| Conduct disorder | Pers. disorder | 0.022 | 0.046 | 0.038 | 0.070 | 0.0065 |

Note. Violent method of death was defined as gun-related, hanging, cutting, or other violent trauma.
